# Characterising Manual Dexterity, Motor Cortex Neuroplasticity and Intracortical Inhibition Long After Burn Injury

**DOI:** 10.1101/2025.10.05.25336730

**Authors:** Grant S. Rowe, Alecia Wood, Mark Fear, Dale W. Edgar, Aleksandra Miljevic, Tyler Osborne, Natalie Morellini, Merrilee Needham, Ann-Maree Vallence, Fiona Wood

## Abstract

**Purpose:** Persistent motor dysfunction after burn injury may be due to altered motor cortex and corticospinal tract function. This study examined manual dexterity, motor cortex neuroplasticity, and intracortical inhibition in former burn patients 1–3 years post-minor injury (<10% TBSA).

**Methods:** Thirty former burn patients (TBSA: 0.78 ± 1.08%) and 30 non-injured controls participated. Manual dexterity was assessed using the Purdue Pegboard. Transcranial magnetic stimulation (TMS) was used to measure motor cortex excitability (motor evoked potential (MEP) amplitude) and short- and long-interval intracortical inhibition (SICI, LICI) before and after paired-associative stimulation (PAS) to induce neuroplasticity.

**Results:** Former burn patients performed worse than controls on the bilateral assembly subtest of the Purdue Pegboard. PAS did not induce changes in MEP amplitude but increased LICI in both former burn patients and control participants. In former burn patients, greater baseline SICI and LICI were associated with better bilateral performance.

**Conclusions:** These findings suggest that poorer motor function persists following minor burn injury and is associated with reduced excitability of motor cortex inhibitory circuits. Further research should explore whether targeting motor cortex inhibitory circuits can improve motor performance following burn injury.

**Highlights:**

- Former burn patients 1–3 years post-minor injury had poorer manual dexterity than controls.
- Paired associative stimulation increased long-interval intracortical inhibition.
- Greater motor cortex inhibition at rest linked to better manual dexterity in former burn patients.

## 1.0 Introduction

Burn injuries can cause extensive damage to the skin and underlying tissues, often leading to motor and sensory dysfunction that significantly diminish quality of life [1]. Motor and sensory dysfunction are evident after both severe burns (>20% TBSA; [2, 3]) and non-severe burns (<20% TBSA; [4, 5]). Indeed, motor dysfunction is commonly observed years after injury in burn survivors, with reports of joint pain, stiffness, gait issues, fatigue, and weakness in the arms and hands, which can impact function in the long-term [2]. The factors contributing to long-term motor dysfunction among burn survivors remain poorly understood [7], even for those with non-severe burn injuries, including minor injuries (<10% TBSA) [8], which is problematic given the significant impact of such injuries on quality of life [9].

The motor cortex and corticospinal tract are essential for voluntary movement, motor learning, and adapting motor functions after injury or disease [10]. It is possible that neuroplasticity—the adaptive capacity of cortical and subcortical networks to reorganise in response to stimuli [11]—may contribute to motor dysfunction after burn injury. Non-severe burns [13], including minor injuries [14], can trigger a systemic inflammatory response that affects the brain [7, 15], which, along with afferent dysfunction [16] and pain [17], may induce changes in cortical function. Therefore, it is possible that motor dysfunction in former burn patients is driven by changes in central nervous system function, particularly in cortical motor areas. Reduced neuroplastic capacity may contribute to persistent challenges in motor control observed in former burn patients. If so, these reductions could represent a potential target for enhancing motor function following burn injuries.

Transcranial magnetic stimulation (TMS) can be used to investigate neuroplasticity [18, 19]. TMS generates electrical currents in the cortex that depolarise neurons in the stimulated cortical region [18]. A suprathreshold TMS pulse delivered to the primary motor cortex (M1) can elicit a motor evoked potential (MEP), measured by electromyography (EMG) from surface electrodes over a target muscle [19, 20]. The peak-to-peak amplitude of the MEP provides a measure of corticospinal excitability, with changes in MEP amplitude providing a marker of neuroplasticity [21]. Administering patterned, repetitive-TMS pulses, such as intermittent theta burst stimulation (iTBS) and continuous theta burst stimulation (cTBS), can modulate corticospinal excitability—iTBS increases excitability, while cTBS decreases excitability—both of which are indicative of neuroplastic changes [22].

Whife et al. [23] used cTBS (two applications of cTBS, known as spaced cTBS) to assess neuroplasticity in the M1 in subacute (<12 weeks) burn patients. A significant association was found between spaced cTBS-induced neuroplasticity 12 weeks post-burn injury and general health perception, as measured by the Short Form-36 (SF36) survey: burn patients who demonstrated the greatest cTBS-induced neuroplasticity reported the highest general health scores. The results provide preliminary evidence that M1 neuroplasticity might be linked to improved functional recovery in burn patients.

GABAergic inhibition, mediated by gamma-aminobutyric acid (GABA), plays a key role in neuroplasticity, particularly in use-dependent adaptations of motor control [24]. It is essential for motor map reorganisation and motor skill acquisition [24], which are important for successful motor function. Given the role of GABAergic inhibition in neuroplasticity, alterations in GABAergic inhibition following burn injury could contribute to persistent challenges in motor control. Understanding these neural mechanisms may provide valuable insights into the long-term impact of burn injuries on motor function and help inform targeted strategies to enhance motor performance in former burn patients.

Motor cortical inhibition can be measured using single- and paired-pulse TMS protocols. Single-pulse TMS delivered during voluntary muscle contraction can be used to measure the cortical silent period (cSP), which reflects the suppression of descending output from M1 [25]. Evidence suggests that the cSP is mediated by GABA_A_ and GABA_B_ receptors at the cortical level [26]. Paired-pulse TMS can be used to measure short- and long-interval intracortical inhibition (SICI, LICI) [19]. For SICI, a subthreshold conditioning stimulus (which is insufficient to elicit an MEP but can modulate neural activity [19, 20]) delivered 1 to 6 milliseconds (ms) before a suprathreshold test stimulus supresses MEP amplitude [27]. For LICI, a suprathreshold conditioning stimulus delivered 100 to 200 ms before a suprathreshold test stimulus also supresses MEP amplitude [28]. Pharmacological evidence suggests that SICI and LICI are mediated by GABA_A_ or GABA_B_ receptor activity, respectively [26, 29–32].

Garside et al. [33] and Whife et al. [34] examined motor cortical inhibition in former and current burn patients using cSP and SICI, respectively. Garside et al. [33] found no significant difference in cSP duration between burn participants and non-injured controls. However, exploratory subgroup analyses (n=13) revealed shorter cSP durations on the burn-affected side in individuals: with upper-limb burns; less than two years post-injury; burns covering <10% TBSA; and partial-thickness burns. These findings suggest that motor cortical inhibition might be reduced in the burn injury population that met these characteristics compared to non-injured controls. In contrast, Whife et al. [34] found no differences in SICI between subacute (6–12 weeks) burn patients and controls, suggesting no change in GABA_A_- mediated intracortical inhibition within the first 12 weeks post-injury. However, given that Garside et al. [33] observed differences in motor cortical inhibition within two years of injury, it could be hypothesised that changes in GABAergic inhibition due to burn injury might occur after the 12-week post-injury period.

Collectively, these studies provide preliminary evidence for changes in neuroplasticity and motor cortical inhibition following burn injuries. Given the prevalence of persistent motor challenges in burn patients, addressing these neural changes is crucial for enhancing motor function and improving long-term outcomes. Thus, the current study aimed to investigate the long-term impact of burn injury on manual dexterity and motor cortex excitability following minor burns (<10% TBSA). We hypothesised that former burn patients would exhibit smaller changes in MEP amplitude following paired associative stimulation (PAS) compared to non-injured controls, indicating a reduced neuroplastic response. Additionally, we expected intracortical inhibition to be lower in the burn group, with greater inhibition correlating with better manual dexterity in both the burn and control groups.

## 2.0 Methods

### 2.1 Participants

Former burn patients were recruited from the Western Australian Adult Burn Unit databases at Fiona Stanley Hospital in Perth, Western Australia; non-injured control participants were recruited from the local community. Inclusion criteria included adult participants (age 18 years or older) with a TBSA injury less than 10%, a burn injury sustained 1–3 years ago, no previous burn injuries, the ability to generate MEPs in response to single-pulse TMS (average MEP amplitude greater than 0.5 mV), and burn types including flame, scald, contact, friction, chemical, and electrical. All participants gave electronic informed consent prior to testing and completed a 17-item safety questionnaire, which was used to screen and exclude individuals with contraindications to TMS [35–37]. The experimental protocol was performed in accordance with the Declaration of Helsinki and was approved by the Murdoch University (2021/047) and South Metropolitan Health Service Human Research Ethics Committee (RGS0000004279).

### 2.2 Experimental Protocol

Figure 1 illustrates the experimental protocol. All participants attended a single 2.5-hour testing session, which included neurophysiological testing as well as motor and sensory function assessments. Additionally, several surveys were sent electronically and completed within 24 hours of the session to provide broader context on self-reported function.

**Figure 1.**
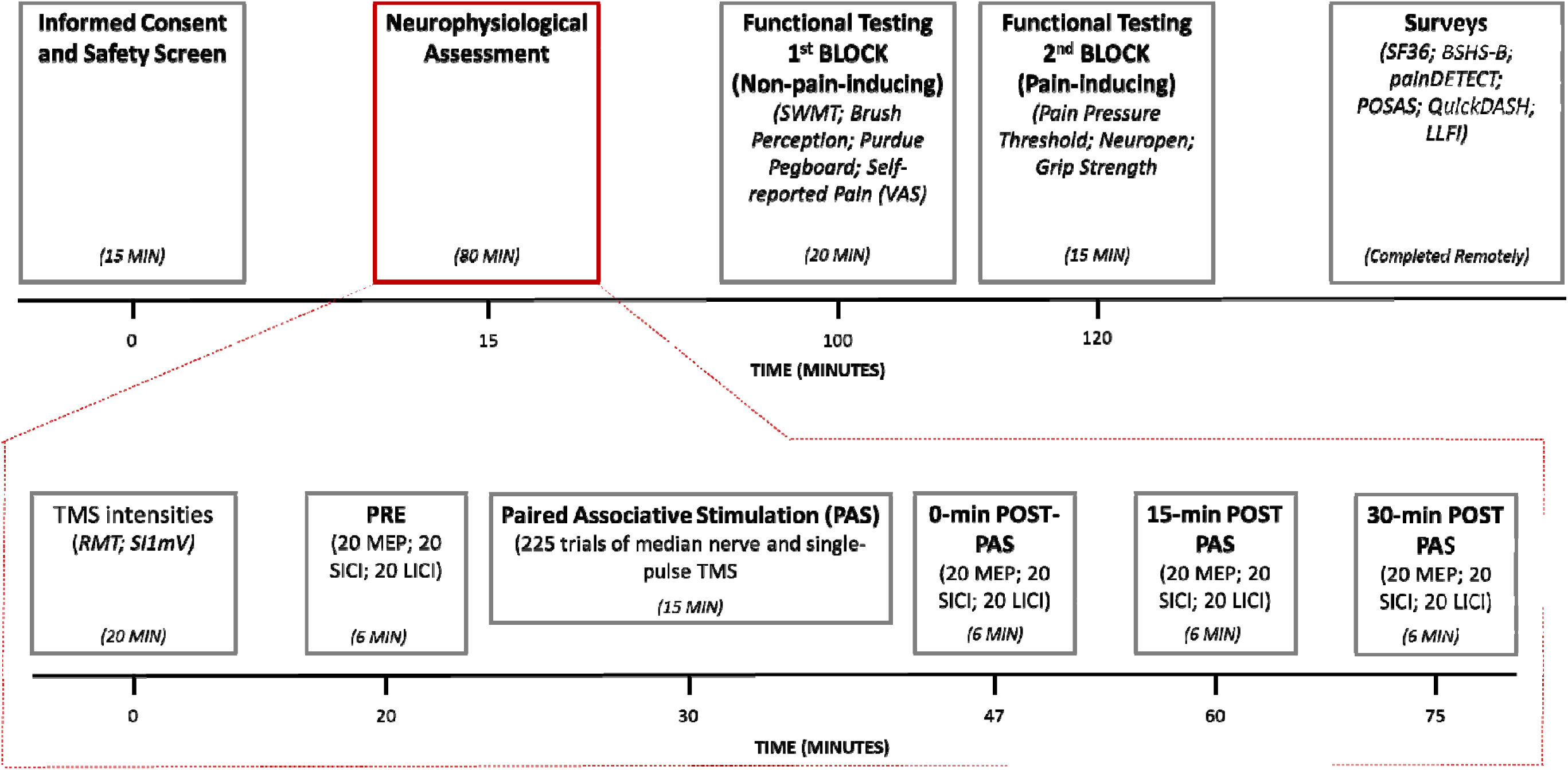
Timeline of the experimental protocol. SWMT: Semmes-Weinstein Monofilament Test; VAS: visual analogue scale; SF36: Short Form-36; BSHS-B: Burn Specific Health Scale-Brief; LLFI: Lower Limb Functional Index; TMS: transcranial magnetic stimulation; RMT: resting motor threshold; SI_1mv_: stimulus intensity required to elicit MEPs with a mean peak-to-peak amplitude of ∼1 mV; MEP: motor evoked potential; SICI: short-interval intracortical inhibition; LICI: long-interval intracortical inhibition.

### 2.3 Functional Testing

#### 2.3.1 Purdue Pegboard

The Purdue pegboard (Lafayette Instrument, USA) was used to examine manual dexterity. Four subtests were included: unilateral dominant hand, unilateral non-dominant hand, simple bilateral, and bilateral assembly. For all subtests, participants were given the opportunity to practice before being instructed to complete the tasks as quickly as possible during testing. For the unilateral subtests, participants were instructed to pick up and place small pegs into holes set out vertically on the pegboard. The testing order for the most-affected and least-affected sides (or the dominant and non-dominant hands for controls) was randomised. The total number of pegs placed within 30 seconds was scored for each hand. For the simple bilateral subtest, participants were instructed to pick up and place the pegs in the holes using both hands simultaneously. The total number of rows (i.e., pairs) with pegs successfully placed in the holes in 30 seconds was scored. For the assembly subtest, participants were instructed to alternate between hands to place and assemble 4-item objects (order of placement: peg, washer, sleeve, and washer) into the holes on the pegboard. The total number of items assembled within 60 seconds was scored.

#### 2.3.2 Short Form-36 (SF36) Health Survey

All participants were asked to complete the Short Form-36 (SF36) [38] remotely via an email link (REDCap Electronic Database, USA). The SF36 has been validated for longitudinal use after burn injury [39]. Eight domain scores were calculated from the survey: physical functioning, role limitations due to physical health, role limitations due to emotional problems, energy/fatigue, emotional well-being, social functioning, pain, and general health [40].

### 2.4 Secondary functional outcomes

Procedure and results for secondary functional outcomes are reported in Supplementary Materials. Specially, surveys including the Burn-specific Health Scale-Brief (S1.1.2), painDETECT (S1.1.3), Patient Observed Scar Assessment Scale (S1.1.4), QuickDASH (S1.1.5), Lower Limb Functional Index (S1.1.6), and sensory functional assessments including the pain pressure threshold (S1.2.1), visual analogue pain scale (S1.2.2), Semmes-Weinstein Monofilament Test (S1.2.3), Neuropen (S1.2.4), and brush perception (S1.2.5). Grip strength was also tested (S1.3).

### 2.5 Neurophysiological Assessment

#### 2.5.1 Electromyography (EMG)

During neurophysiological testing, participants sat on a chair with their arms supported by a pillow. EMG activity was recorded using Ag-AgCI surface electrodes placed in a belly-tendon montage on the first dorsal interosseous (FDI) muscle [41]. EMG activity was amplified (x1000) using a CED 1902, bandpass filtered (20-1000 Hz) and digitised at 5 kHz using a CED 1401 (Cambridge Electronic Design, UK). In burn participants, EMG activity was recorded from the hand on the burn affected side of the body; if the burn injury affected both sides of the body, the more affected side was determined based on burn surface area and depth and used as the test side. The testing side for control participants was closely matched to that of the burn participants. In the burn group, 20 participants were tested on their left dominant hemisphere (right-handed), 9 on their right non-dominant hemisphere (right-handed), and 1 on their left non-dominant hemisphere (left-handed). In the control group, 19 participants were tested on their left dominant hemisphere (right-handed), 10 on their right non-dominant hemisphere (right-handed), and 1 on their left non-dominant hemisphere (left-handed).

#### 2.5.2 Transcranial Magnetic Stimulation (TMS)

Two MagStim 200^2^ stimulators connected by a MagStim BiStim module (Magstim, UK) were used to generate single and paired pulses. Monophasic pulses were delivered through a figure-of-eight coil (90-mm diameter) placed tangentially to the scalp with the coil handle positioned backwards and rotated away from the midline by ∼45° to induce a posterior-anterior current in M1 contralateral to the most-affected hand. The optimal site of M1 stimulation was defined as the site that elicited the largest and most consistent MEPs in the FDI muscle [37, 42]. The optimal site was marked on a tight-fitting skull cap to ensure reliable placement of the coil throughout the testing session. Resting motor threshold (RMT) was defined as the minimum stimulus intensity (as a percentage of the maximum stimulator output: MSO) required to elicit MEPs ≥ 50 µV in at least three out of five consecutive trials [37]. SI_1mV_ was defined as the stimulus intensity (as a percentage of MSO) required to elicit MEPs with a mean peak-to-peak amplitude of ∼1 mV. All measures were obtained with the target muscle at rest.

#### 2.5.3 Intracortical Inhibition

Short-interval intracortical inhibition (SICI) and long-interval intracortical inhibition (LICI) were assessed by delivering single- and paired-pulse stimuli. For single-pulse trials, a test stimulus at SI_1mV_ was delivered. For SICI paired-pulse trials, a conditioning stimulus at 80% RMT was delivered before a test stimulus at SI_1mV_, with an inter-stimulus interval of 3 ms [43]. For LICI paired-pulse trials, a conditioning stimulus at SI_1mV_ was delivered before a test stimulus at SI_1mV_, with an inter-stimulus interval of 100 ms [44]. Signal software (CED, UK) was used to pseudo-randomise trial conditions (SI_1mV_-alone, SICI, and LICI trials) with an inter-trial interval of 5 s (±20%). Single- and paired-pulse trials for SICI and LICI were delivered in blocks, with each block comprising 60 trials: 20 SI_1mV_-alone, 20 SICI, and 20 LICI (total block time ∼6 minutes). Participants were instructed to remain quiet, keep their eyes open, and stay attentive but relaxed during TMS testing.

#### 2.5.4 Paired Associative Stimulation (PAS)

Paired associative stimulation (PAS) was chosen to examine neuroplasticity responses because it induces long-term potentiation (LTP)-like plasticity, driven by the Hebbian principle of spike-timing-dependent plasticity [45]. This capacity for neuroplasticity, particularly through sensory-motor integration, was selected for the burn population, as sensory dysfunction can significantly impact motor function and long-term functional outcomes [46, 47]. PAS was delivered by pairing median nerve stimulation on the most-affected side with single-pulse TMS to the contralateral M1 [48]. Peripheral nerve stimulation was administered with 200 microsecond (µs) square width pulses with a direct current stimulator (Digitimer [DS7], Digitimer, UK), with bipolar surface electrodes: the anode was placed over the median nerve at the wrist and the cathode was placed 3 cm proximal to the anode. Sensory perceptual threshold was determined using a stepwise procedure with two reversals. The peripheral nerve stimulation intensity for PAS was set to 3x the sensory perceptual threshold, the TMS pulses were applied to the contralateral M1 using a single Magstim 200^2^ at SI_1mV_ (obtained from the single Magstim 200^2^), and the interval between nerve stimulation and TMS was 25 ms. PAS consisted of 225 trials of paired median nerve and single-pulse TMS applied at a rate of 0.25 Hz for 15 mins [45]. To direct attention during PAS, a light emitting diode (LED) was placed in front of the participants target hand and randomly flashed in 20-40 trials throughout the PAS protocol (Signal software): participants were instructed to attend to the hand throughout PAS and report the number of LED flashes after PAS [49]. The mean absolute error between reported and true counts was 2.4 ± 3.2 in former burn patients and 2.3 ± 4.3 in controls. No participants were excluded from the analysis due to LED counting errors.

Single-pulse MEP amplitude, SICI, and LICI, were assessed at four time points: before PAS (PRE), immediately after PAS (0-min POST-PAS), 15 minutes post-PAS (15-min POST-PAS), and 30 minutes post-PAS (30-min POST-PAS) (see Figure 1). Participants also rated their self-perceived alertness/sleepiness using the 10-step Karolinska Sleepiness scale after each testing block, including after PAS [50]. Self-reported alertness/sleepiness scores increased from baseline (PRE) to immediately post-PAS in both former burn patients (4.7 ± 1.7 to 5.7 ± 1.7) and controls (4.6 ± 1.9 to 5.6 ± 1.8). Scores then gradually declined across subsequent PAS blocks, returning close to baseline levels by 30-min POST-PAS (former burn patients = 4.9 ± 1.5; controls = 5.4 ± 1.2).

Baseline (PRE) SICI results were also examined in former burn patients exhibiting atypical facilitation, defined as a SICI ratio exceeding 1.5. Atypical facilitation in SICI protocols is linked to poorer manual dexterity and is considered distinct from individuals with reduced SICI, whose ratio values typically fall below or around 1.0 [51]. A clinical summary of burn participants identified as displaying atypical facilitation is provided in the results section.

### 2.6 Data Processing

All single- and paired-pulse TMS trials were recorded using Signal data acquisition and analysis software (CED, UK). A customised macro was used to analyse trials, determining MEP peak-to-peak amplitudes from 10 to 60 ms (and 110 to 160 ms for LICI) after the stimulus and background EMG activity 100 ms before the stimulus artifact (root mean square [RMS]). Frames were excluded if the background EMG activity exceeded the RMS mean + (2x SD) across all session frames. The proportion of trials removed due to excessive EMG background noise (mean ± SD) was 2.0% ± 1.2 in former burn patients and 2.2% ± 1.7 in controls.

### 2.7 Data Analysis

Data analysis was performed using R Statistical software (version 4.4.1) [52]. A generalised linear mixed model (GLMM) [R: lme4] [53] with a gamma distribution family (log) was used to explore the interaction effects between GROUP (former burn patients, non-injured controls), TIME (PRE, 0-min POST-PAS, 15-min POST-PAS, 30-min POST-PAS), and TMS TYPE (SI_1mV_-alone, SICI, LICI) on MEP amplitude trial data. The random effect structure included participant random intercepts and slopes for GROUP, TIME, and TMS TYPE. Another GLMM analysis was conducted with only the baseline (PRE) data, with the random effect structure including participant random intercepts and slopes for GROUP and TMS TYPE. GLMMs were chosen because they allow for the analysis of complex, repeated measures data by accounting for both fixed and random effects, handling variability across participants and experimental groups, and providing the flexibility to select the appropriate family distribution to match the data. Model fitting was assessed by comparing nested models using Akaike Information Criterion (AIC) and chi-square likelihood ratio tests [R: lme4] [53]. AIC was used to evaluate the trade-off between goodness of fit and model complexity, while the chi-square test determined whether additional parameters significantly improved model fit. Wald Chi-Squared tests were used for null hypothesis significance testing of main and interaction effects in GLMM. Post-hoc estimated marginal means analyses [R: emmeans] [54] with Holm-Bonferroni adjustment for multiple comparisons were conducted for significant interaction effects (*p* < 0.05). The pegboard tests, baseline TMS intensity data, and SF36 responses were analysed using independent samples t-tests with non-parametric bootstrapping (1,000 simulations) [R: boot] [55], with significance detected when the Holm-Bonferroni adjusted *p*-values were < 0.05. Non-parametric bootstrapping enhances independent sample t-test analysis by providing a robust method to estimate sampling distributions, improving the accuracy of confidence intervals (CIs) and *p*-value estimation. This approach is particularly useful when data assumptions (e.g., normality) are not met or when dealing with unequal sample sizes. Cohen’s *d* effect size was calculated for pegboard scores to quantify the magnitude of difference between the burn and control groups, providing insight into the practical significance of the results. This was followed by bootstrapping (1,000 simulations) [R: boot] [55] to estimate the 95% CI for the effect sizes. Confidence intervals that cross zero indicate a non-significant difference between effect sizes, suggesting comparable magnitudes of effect across pegboard subtests. Exploratory correlation analyses between bilateral pegboard scores, MEP amplitude changes following PAS, and PRE intracortical inhibitory outcomes were performed using Pearson’s correlation coefficient (*r*) with non-parametric bootstrapping (1,000 simulations) [R: boot] [55]. Significance was determined if the 95% CI of the correlation coefficient did not cross zero. To assess whether the strength of the correlations differed between groups, bootstrapping was performed by resampling the data and calculating the difference between the correlation coefficients at each iteration. A significant difference was identified if the resulting 95% CI for the differences did not cross zero. Significance in exploratory correlation analyses was determined based on overlapping CIs rather than *p*-values to account for uncertainty in effect estimates and to provide a more robust interpretation of the relationships. Correlation analyses were limited to the bilateral pegboard subtests, as TMS testing was conducted on only one hemisphere, allowing the full sample to be included in the analyses. Data preprocessing and visualisation were performed using the following R packages: dplyr [56], ggplot2 [57], ggsignif [58], cowplot [59], grid [52], and gridExtra [60]. The primary data are available via https://osf.io/3xfkp/?view_only=48fa8a1f1b1c473d9e7f4748f61ecaf0. Data analysis methods for secondary outcomes is provided in relevant section in Supplementary Materials.

## 3.0 Results

### 3.1 Summary of Clinical Characteristics for Former Burn Patients

Thirty former burn patients (1–3 years post-injury) and 30 non-injured control participants took part in the study. The two groups were closely matched for sex (burn group: females = 13; control group = 16), age (burn group: 43.6 ± 14.0 years; control group: 43.3 ± 14.6 years), and dominant hemisphere tested (burn group: 20; control group: 19). In the burn group, the average time since injury was 82 ± 17 weeks, with a mean TBSA affected of 0.78 ± 1.08%. The distribution of burn types included contact (n=12), scald (n=11), flame (n=6), and low-voltage electrical (n=1). Burn depth classifications were superficial (n=10), partial-thickness (n=12), deep (n=6), and full-thickness (n=2). Only two-thirds (n=21) of the injuries involved the upper limb, though these did not physically prevent task execution.

### 3.2 Pegboard Performance

Figure 2 shows pegboard performance for the four subtests in former burn patients (1–3 years post-minor injury) and non-injured controls. Bootstrapped t-tests showed that burn participants demonstrated poorer performance compared to controls for the assembly subtest (95% CI: −7.95 to −1.63, *p* = 0.016, *d* = −0.78). However, no significant differences were observed between groups for the dominant hand subtest (95% CI: −1.92 to 0.01, *p* = 0.140, *d* = −0.52), the non-dominant hand subtest (95% CI: −1.52 to 0.39, *p* = 0.242, *d* = −0.31), or the simple bilateral subtest (95% CI: −1.61 to −0.03, *p* = 0.140, *d* = −0.53). Effect size differences across pegboard subtests were non-significant, indicating comparable group differences across all measures. The largest contrast—between assembly (*d* = −0.78) and non-dominant hand (*d* = −0.31)—did not reach statistical significance (95% CI: −1.09 to 0.13). Given the consistency of effect sizes across subtests and their alignment with the assembly task—where burn participants consistently performed worse than controls—these findings suggest a broadly reduced motor capacity rather than a deficit specific to individual tasks.

**Figure 2.**
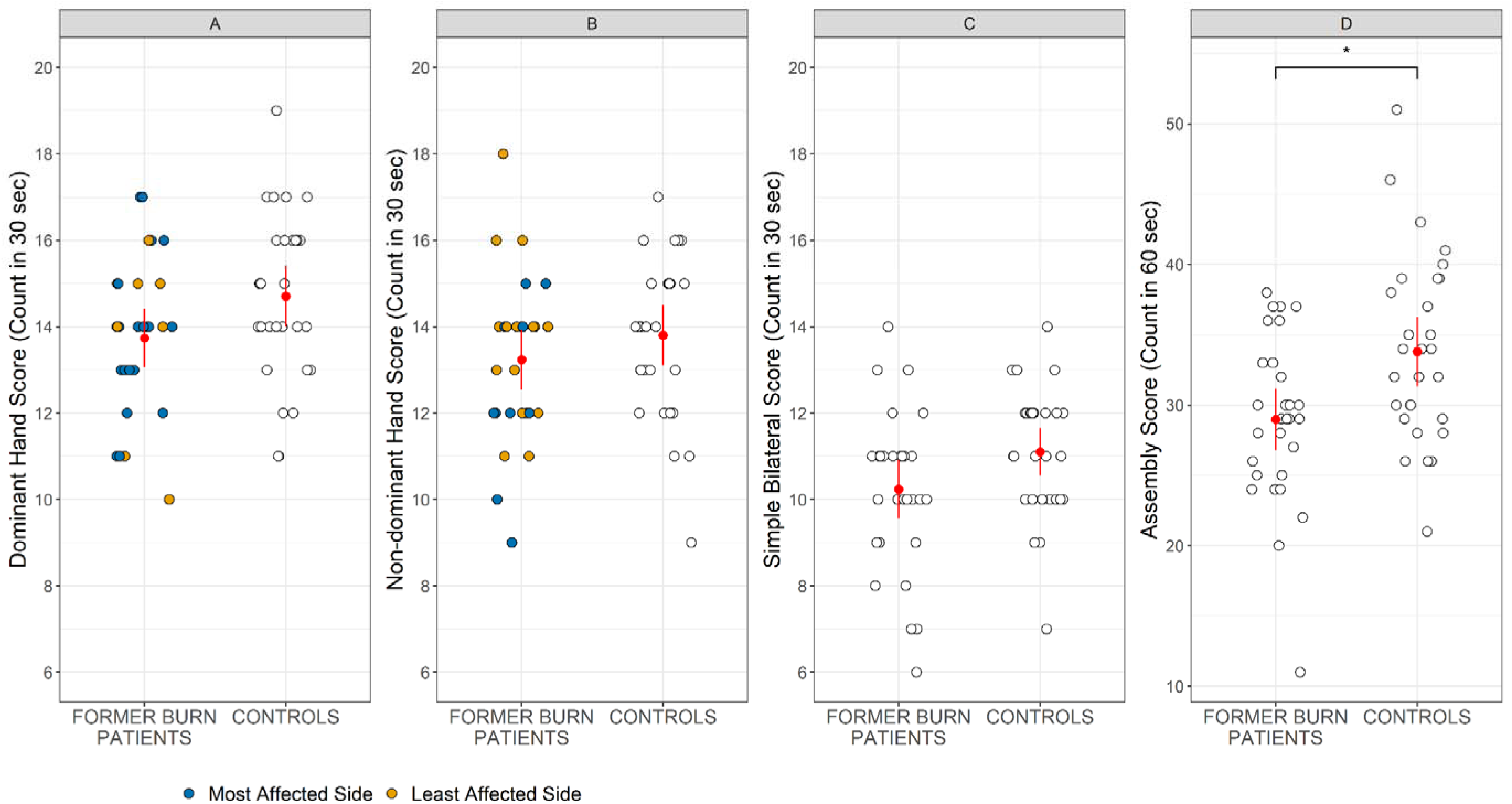
(A) Dominant hand, (B) non-dominant hand, (C) simple bilateral, and (D) assembly pegboard performance in former burn patients (1-3 years post-minor injury) and non-injured controls. Red symbols represent the mean, with error bars indicating confidence intervals (CI); other symbols represent individual participants. An asterisk indicates a statistically significant difference between groups

### 3.3 Neurophysiological assessment

#### 3.3.1 Baseline TMS Intensities (RMT; SI_1mV_)

The average RMT and SI_1mV_ in former burn patients (1–3 years post-minor injury) were 50.9 ± 6.9% (MSO) and 59.6 ± 8.9% (MSO), respectively. In non-injured controls, the average RMT and SI_1mV_ were 50.4 ± 7.8% (MSO) for RMT and 58.4 ± 8.8% (MSO). Bootstrapped t-tests showed no significant differences between burn participants or non-injured controls for RMT (95% CI: −3.39 to 4.02, *p* = 1.000) or SI_1mV_ (95% CI: −3.16 to 5.25, *p* = 1.000). These findings indicate that corticospinal excitability, as measured by RMT and SI_1mV_, was comparable between burn and control groups, suggesting no significant differences in baseline motor cortex excitability.

#### 3.3.2 MEP Amplitude, SICI, and LICI Following PAS

Figure 3 presents MEP amplitude, while Figure 4 displays SICI and LICI before and after PAS in former burn patients (1–3 years post-minor injury) and non-injured controls. A GLMM was used to evaluate changes in MEP amplitude elicited by single-pulse trials, paired-pulse SICI trials, and paired-pulse LICI trials following PAS in the burn and control groups. There were no significant effects for GROUP (χ2 (N = 60) = 1.76, *p* = 0.185) or TIME (χ2 (N = 60) = 0.71, *p* = 0.870). However, there was a significant main effect for TMS TYPE (χ2 (N = 60) = 131.03, *p* < 0.001) and a significant TIME x TMS TYPE interaction (χ2 (N = 60) = 62.09, *p* < 0.001). No additional significant interaction effects were observed, including those related to GROUP.

**Figure 3.**
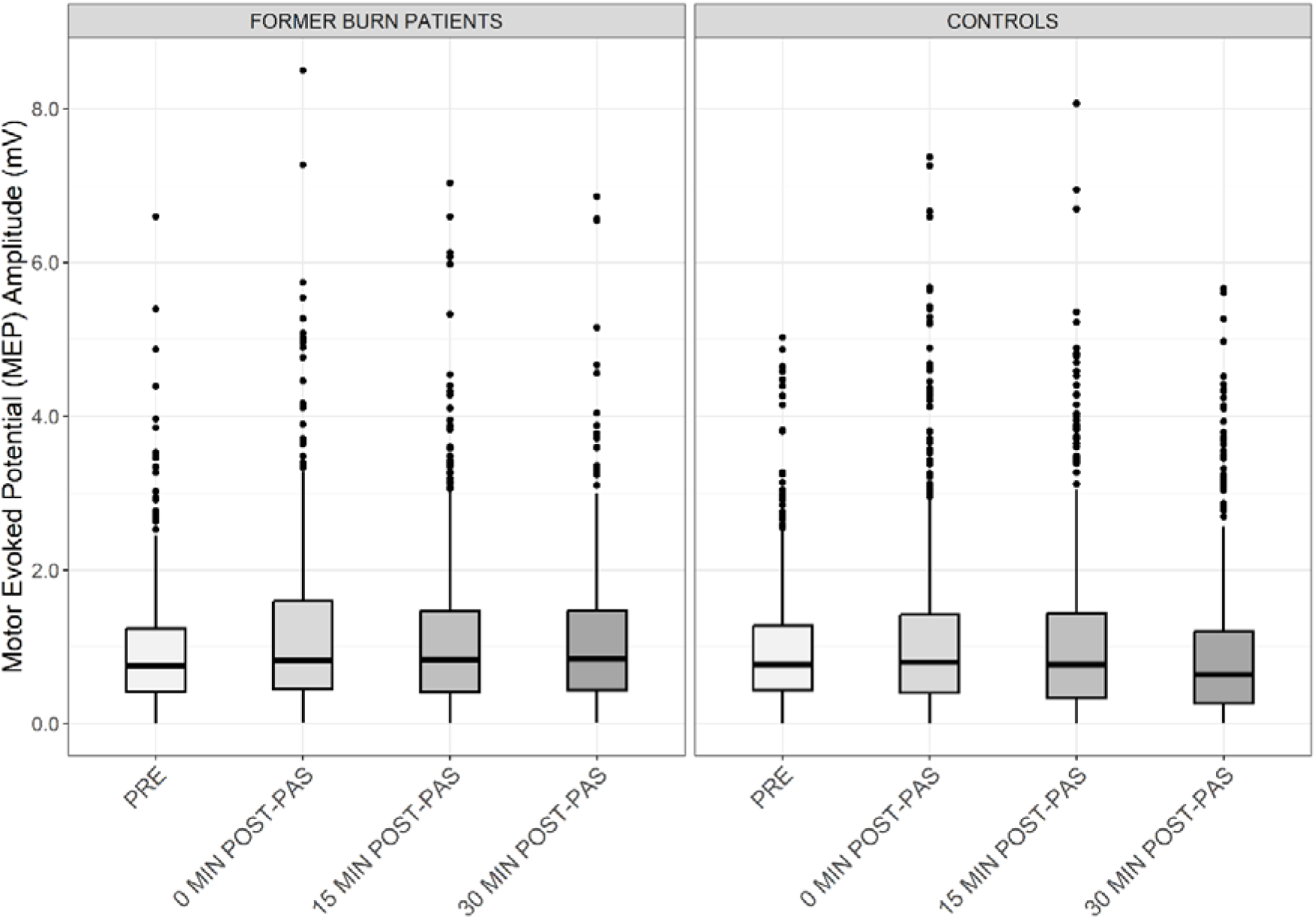
Single-pulse MEP amplitude following PAS in former burn patients (1-3 years post-minor injury) (left panel) and non-injured controls (right panel). The lower and upper hinges represent the 25th and 75th percentiles, respectively. The whiskers extend to values that are up to 1.5 times the interquartile range (the difference between the 25th and 75th percentiles) below the lower hinge and above the upper hinge. Points that lie beyond the whiskers are shown as filled circles, indicating trial-level outliers.

**Figure 4.**
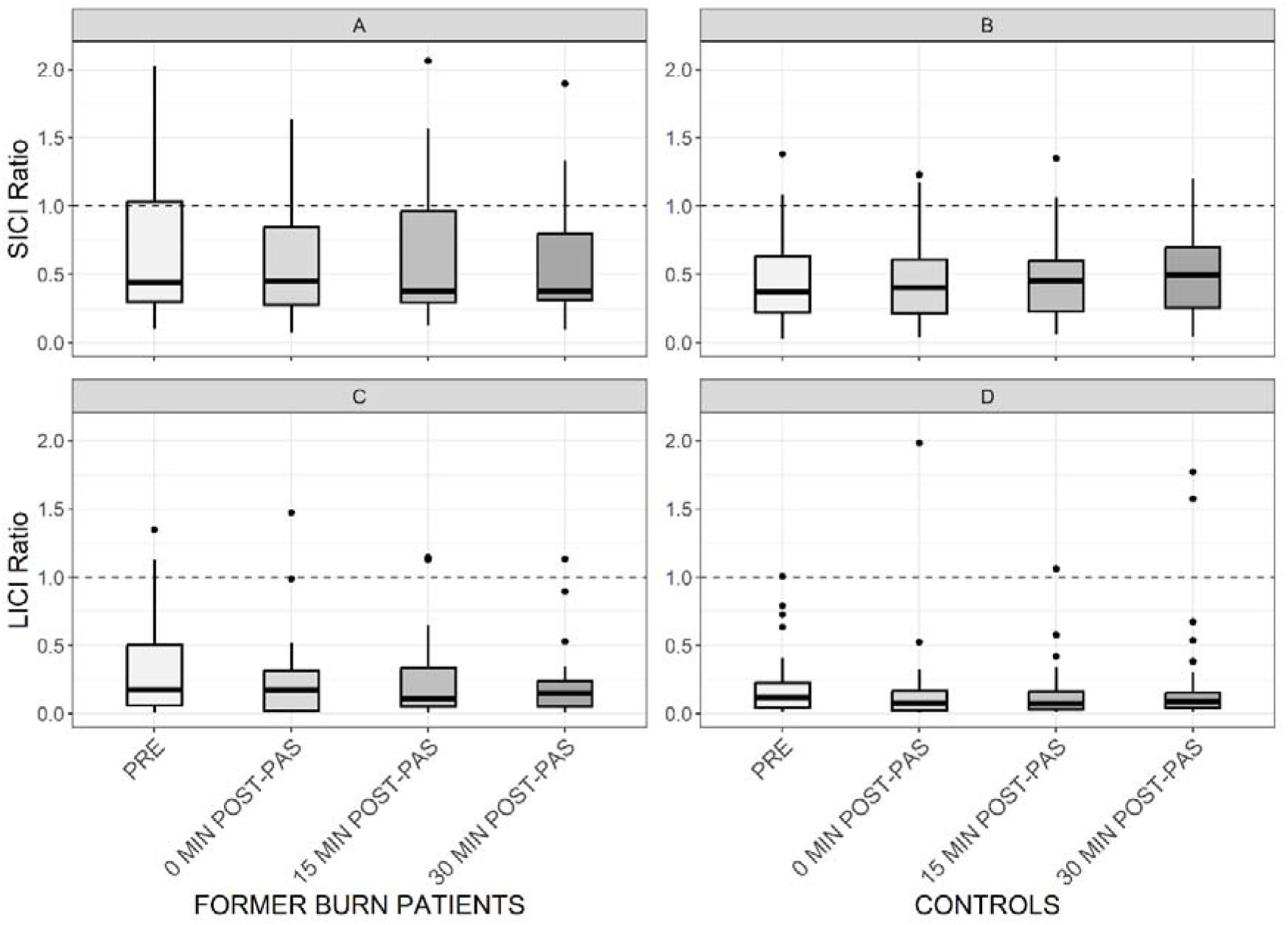
Short-interval intracortical inhibition (SICI) and long-interval intracortical inhibition (LICI) ratios following PAS in former burn patients (1-3 years post-minor injury) (panels A and C, respectively) and non-injured controls (panels B and D, respectively). Points that lie beyond the whiskers are shown as filled circles, indicating aggregated-level outliers. The y-axis has been cropped at 2.1 to highlight the boxplots (see Supplementary Material for uncropped version: section 1.4.1 and Figure S6).

Post hoc analyses showed no significant changes in single-pulse MEP amplitude (*z* < 1.95, *p* > 0.304) or SICI (*z* < 2.01, *p* > 0.152) across TIME, but LICI showed a significant increase from PRE to 0-min POST-PAS (*z* = −5.96, *p* < 0.001) and 15-min POST-PAS (*z* = −4.56, *p* < 0.001). These findings indicate that while PAS had no significant effect on MEP amplitude or SICI, it led to a transient increase in LICI, suggesting a selective modulation of long-acting intracortical inhibition.

Bootstrapped correlation analyses examined the relationships between changes in MEP amplitude and bilateral pegboard performance. No significant associations were found between MEP amplitude changes at 0-min POST-PAS and performance on the bilateral pegboard subtests in either former burn patients (1–3 years post-minor injury) or non-injured controls, suggesting no clear link between PAS-induced neuroplasticity and motor performance. These results are presented in Supplementary Materials (see section S1.4.2 and Figure S7).

#### 3.3.3 Baseline (PRE) SICI and LICI Comparison Between Former Burn Patients and Non-injured Controls

A GLMM was used to evaluate baseline (PRE) differences in SICI and LICI between former burn patients (1–3 years post-minor injury) and non-injured controls. There were no significant effects for GROUP (χ2 (N = 60) = 2.14, *p* = 0.143) and no significant GROUP x TMS TYPE interaction (χ2 (N = 60) = 4.03, *p* = 0.133). However, there was a significant main effect of TMS TYPE (χ2 (N = 60) = 138.78, *p* < 0.001). LICI ratios were numerically smaller than SICI ratios (Figure 4). These findings indicate no significant differences in baseline SICI and LICI between burn and control groups.

#### 3.3.4 Associations Between Baseline SICI and LICI and Bilateral Pegboard Performance

Figure 5 presents scatterplots depicting the associations between SICI magnitude and bilateral pegboard performance. Exploratory bootstrapped correlation analyses were conducted to examine the relationships between baseline intracortical inhibition and bilateral pegboard scores. In former burn patients (1–3 years post-minor injury), baseline SICI was significantly negatively correlated with simple bilateral subtest performance (*r* = −0.26, 95% CI: −0.54 to −0.01), indicating that greater baseline SICI was associated with better manual dexterity (Figure 5A). However, baseline SICI was not significantly associated with assembly subtest performance in burn participants (*r* = −0.31, 95% CI: −0.62 to 0.05) (Figure 5B). There were no significant associations observed between baseline SICI and performance on the bilateral subtests in non-injured controls.

**Figure 5.**
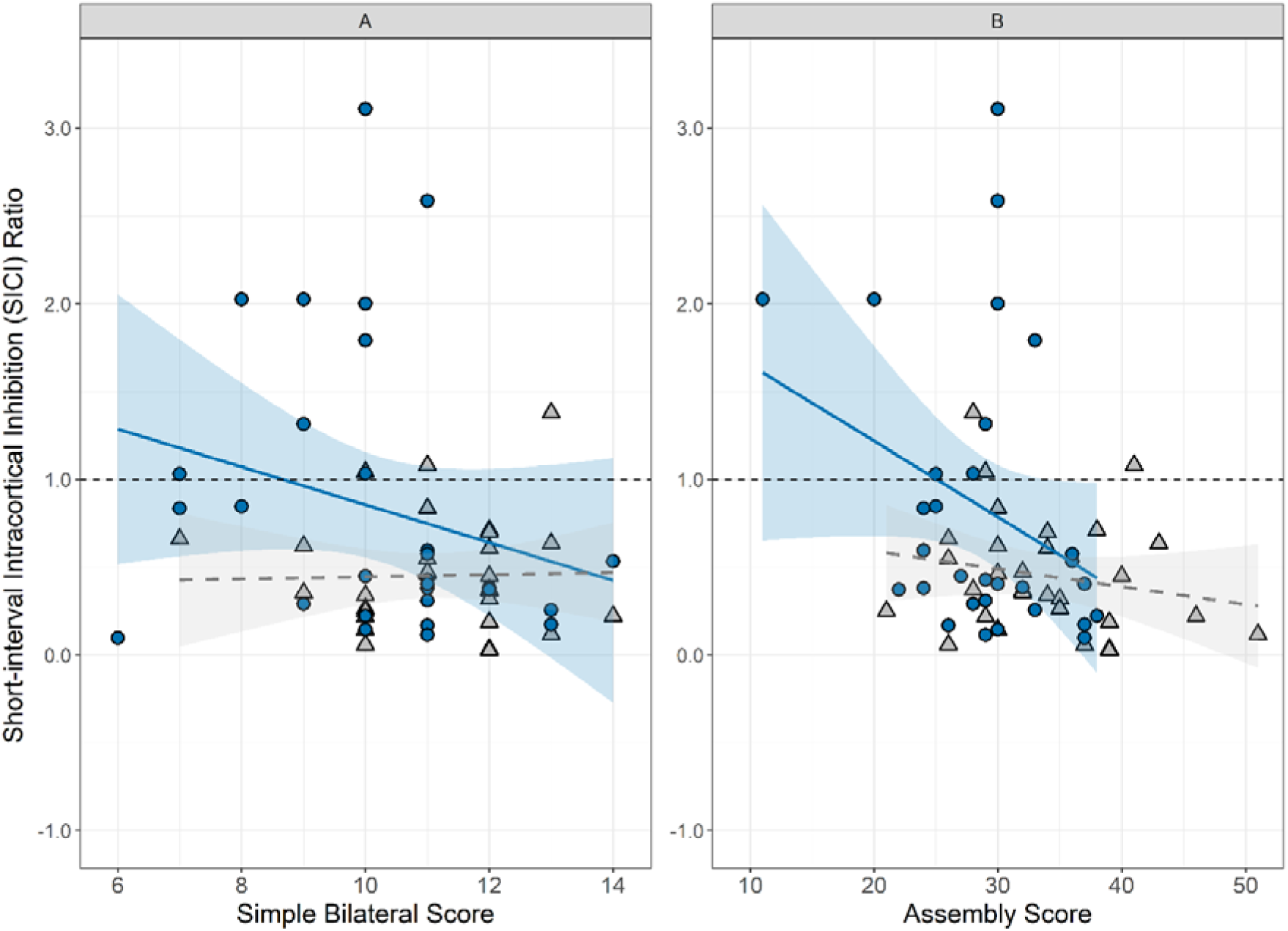
The relationships between the magnitude of SICI and bilateral pegboard performance in former burn patients (1-3 years post-minor injury) (blue circle symbols) and controls (grey triangle symbols). Exploratory analyses revealed that greater baseline SICI is associated with better manual dexterity for the simple bilateral subtest in burn participants.

Figure 6 presents scatterplots illustrating the associations between LICI magnitude and bilateral pegboard performance. In former burn patients (1–3 years post-minor injury), baseline LICI showed significant negative correlations with simple bilateral (*r* = −0.43, 95% CI: −0.67 to −0.17) and assembly subtest performance (*r* = −0.28, 95% CI: −0.51 to −0.01), indicating that greater baseline LICI was associated with better manual dexterity (Figure 6A, B). In non-injured controls, baseline LICI was significantly positively correlated with simple bilateral subtest performance (*r* = 0.31, 95% CI: 0.06 to 0.54), suggesting that lower baseline LICI was associated with better manual dexterity (Figure 6A). However, baseline LICI was not significantly associated with assembly subtest performance in non-injured controls (*r* = 0.40, 95% CI: −0.04 to 0.68) (Figure 6B).

**Figure 6.**
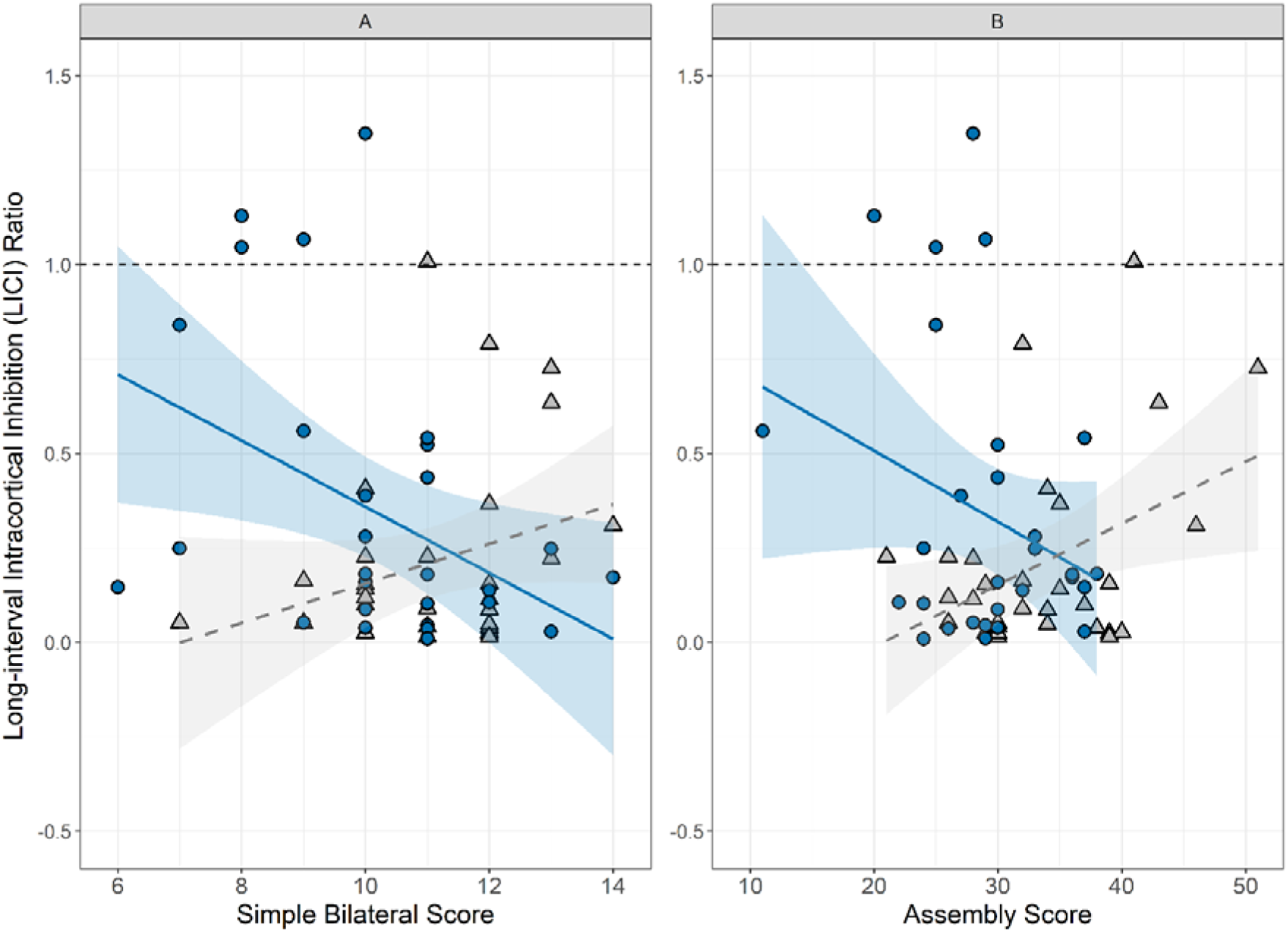
The relationships between the magnitude of LICI and bilateral pegboard performance in former burn patients (1-3 years post-minor injury) (blue circle symbols) and controls (grey triangle symbols). Significant differences were observed between groups in the relationship patterns between baseline LICI and bilateral pegboard subtests.

Bootstrapped correlation analyses were performed to examine group differences in the relationships between baseline inhibition and motor performance. Significant differences emerged between former burn patients (1–3 years post-minor injury) and non-injured controls in the associations between baseline LICI and bilateral pegboard subtests. Specifically, significant group differences were observed for the simple bilateral (95% CI: −1.06 to −0.38) and assembly subtests (95% CI: −1.04 to −0.20). These findings suggest that the relationship between GABA_B_-mediated intracortical inhibition and manual dexterity differs between former burn patients and non-injured controls, highlighting potential differences in the role of intracortical inhibition in motor function.

### 3.4 Characteristics of Former Burn Patients Who Showed Atypical SICI Facilitation

Six former burn patients (1–3 years post-minor injury) exhibited atypical facilitation to the paired-pulse SICI protocol at baseline (PRE), defined by a SICI ratio exceeding 1.5. No distinct clinical characteristics were identified in these atypical facilitators. The clinical profile of this atypical facilitation subgroup generally resembled that of the full sample: average time since injury of 89 weeks (min–max: 70–124), with burn types including contact (n=3), scald (n=1), flame (n=1), and electrical (n=1), and burn depths including superficial (n=3), partial-thickness (n=1), deep (n=1), and full-thickness (n=1). The mean TBSA affected in this atypical facilitation subgroup was 0.42% (min–max: 0.01–2.00). The atypical facilitation subgroup included two individuals tested on their non-dominant hemisphere and compared to the full sample, was numerically older, with a mean age of 54.2 years (min– max: 48–63).

### 3.5 Short Form-36 (SF36)

Table 1 presents the mean and standard deviation of SF36 domain scores, along with statistical significance results. Figure S1 shows SF36 domain scores for role limitations due to physical health in former burn patients (1–3 years post-minor injury) and controls (see Supplementary Materials: 1.1.1). Missing values in the SF36 survey (n=1, a control participant who did not submit the survey) were handled by excluding incomplete cases from the analysis. Bootstrapped t-tests revealed no significant differences between burn participants and non-injured controls across all SF36 domain scores, including role limitations due to physical health, after *p*-value adjustment (burn group: 86.7 ± 21.3, control group: 95.5 ± 11.4, 95% CI: −17.8 to −0.7, *p* = 0.425). These results suggest that former burn patients’ overall health and well-being are not significantly different from those of non-injured controls. Importantly, 17 burn participants and 21 controls reported the maximum score of 100 for role limitations due to physical health, reflecting a ceiling effect, which was consistently observed across all SF36 domains. This reduced variability in bootstrap samples, potentially narrowing confidence intervals and underestimating uncertainty. The ceiling effect should be considered when interpreting the results.

**Table 1:**
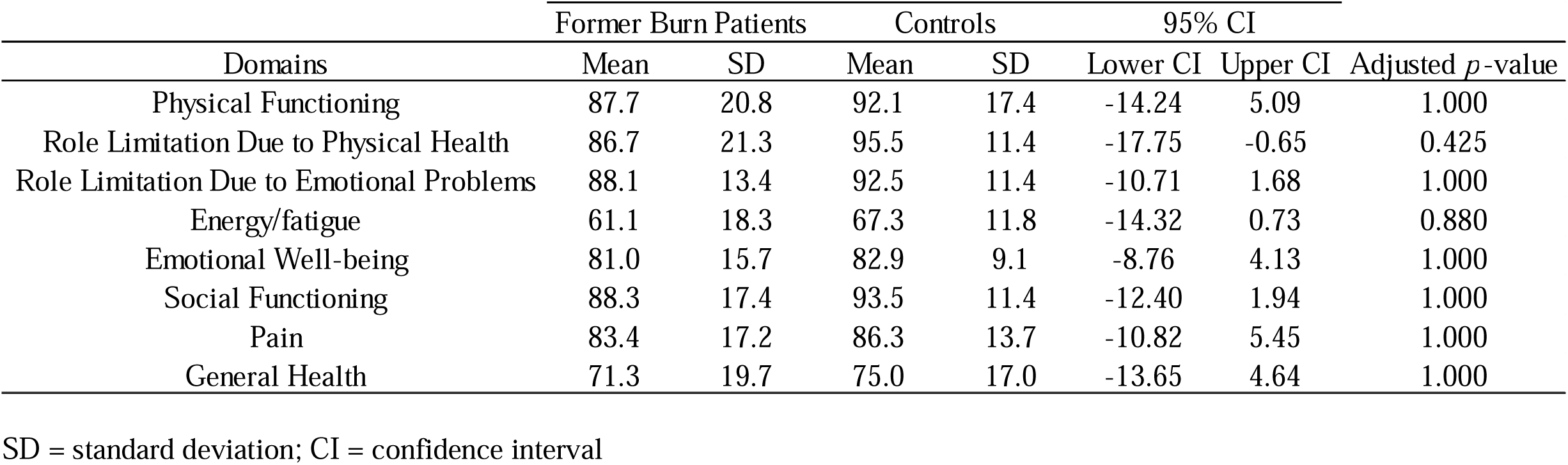
Short Form-36 domain scores in former burn patients (1–3 years post-minor injury) (n=30) and non-injured controls (n=29).

| Domains | Former Burn Patients |  | Controls |  | 95% CI |  | Adjusted *p*-value |
| --- | --- | --- | --- | --- | --- | --- | --- |
|  | Mean | SD | Mean | SD | Lower CI | Upper CI |  |
| Physical Functioning | 87.7 | 20.8 | 92.1 | 17.4 | -14.24 | 5.09 | 1.000 |
| Role Limitation Due to Physical Health | 86.7 | 21.3 | 95.5 | 11.4 | -17.75 | -0.65 | 0.425 |
| Role Limitation Due to Emotional Problems | 88.1 | 13.4 | 92.5 | 11.4 | -10.71 | 1.68 | 1.000 |
| Energy/fatigue | 61.1 | 18.3 | 67.3 | 11.8 | -14.32 | 0.73 | 0.880 |
| Emotional Well-being | 81.0 | 15.7 | 82.9 | 9.1 | -8.76 | 4.13 | 1.000 |
| Social Functioning | 88.3 | 17.4 | 93.5 | 11.4 | -12.40 | 1.94 | 1.000 |
| Pain | 83.4 | 17.2 | 86.3 | 13.7 | -10.82 | 5.45 | 1.000 |
| General Health | 71.3 | 19.7 | 75.0 | 17.0 | -13.65 | 4.64 | 1.000 |

SD = standard deviation; CI = confidence interval

## 4.0 Discussion

This study showed that former burn patients (1–3 years post-minor injury) exhibited poorer bilateral assembly subtest performance than non-injured controls, as assessed by the Purdue Pegboard test. PAS led to an increase in LICI but no change in MEP amplitude or SICI in either burn or control groups. There were no differences in baseline SICI or LICI between burn participants and non-injured controls, however, exploratory analyses showed that greater baseline SICI and LICI were associated with better manual dexterity in former burn patients but not non-injured controls.

### Poorer Manual Dexterity in Former Burn Patients Compared to Controls Persists for 1–3 Years Following Injury

Compared to non-injured controls, former burn patients (1–3 years post-minor injury) demonstrated poorer performance on the bilateral assembly Purdue Pegboard subtest (Figure 2). This finding, in former burn patients with minor injuries (<10% TBSA), is consistent with previous research showing self-reported motor dysfunction (measured using the QuickDASH) in individuals with non-severe burn injuries [4, 61]. Furthermore, these results provide objective evidence of reduced manual dexterity in former burn patients compared to controls, persisting for at least one year post-injury. Although effect size differences across the unilateral and simple bilateral Purdue Pegboard subtests were non-significant, results followed a similar trend to the assembly subtest—poorer Purdue Pegboard performance in former burn patients than in non-injured controls—further supporting a persistent reduction in manual dexterity in minor burn injury populations. It is interesting to note that the assembly subtest requires greater cognitive effort than the other pegboard subtests, as evidenced by significantly higher activation in the left prefrontal cortex detected via functional Near-Infrared Spectroscopy (fNIRS) [62]. The greater cognitive effort may exacerbate the reduced manual dexterity in former burn patients and partially account for the significant performance difference between burn participants and controls on the assembly subtest, but not on the less complex subtests.

Notably, poorer manual dexterity was observed across a diverse sample of burn participants, including nine with face, trunk, or lower-limb injuries, as well as within the subgroup of those with upper-limb burns (see Supplementary Materials section 2.2 and Figure S10 for subgroup analysis of upper-limb burn injury participants). These findings support the notion that burn injuries have widespread, systemic effects on the neural and musculoskeletal systems that are involved in motor control [for review see: 7, 15]. Additionally, given that this study recruited former burn patients with minor injuries, behavioural adaptations—such as movement compensations—may have contributed to the diminished performance, underscoring the complex interplay between injury and functional adaptation [63].

### No PAS-induced Neuroplasticity in Burn Injury or Non-injured Control Participants

The current study found no change in MEP amplitude following PAS in either former burn participants (1–3 years post-minor injury) or non-injured controls (Figure 3). Although PAS is thought to induce LTP-like plasticity through sensory-motor integration [48]—a key process for burn populations given the impact of sensory dysfunction on motor control and recovery [46, 47]—the present findings do not show any evidence of PAS-induced LTP-like plasticity in either group. To the best of our knowledge, only one previous study has examined M1 plasticity following burn injury. The previous study showed that, compared to controls, subacute burn patients exhibited reduced long-term depression (LTD)-like plasticity induced by spaced continuous theta burst stimulation (cTBS) at 6 weeks post-injury, but not at 12 weeks post-injury [23]. This previous finding suggests a reduced capacity for LTD-like neuroplasticity in the subacute period following injury (i.e., ∼6 weeks), which normalises later in recovery (i.e., ∼12 weeks). In light of the current findings, which show no LTP-like plasticity in former burn patients (1–3 years post-minor injury), it is plausible that changes in neuroplasticity have normalised long after the injury. Alternatively, it could suggest that burn injury primarily affects LTD-like plasticity, induced by cTBS, rather than LTP-like plasticity, typically induced by PAS, as these mechanisms differ in how they influence neural responses. It is also important to note that the PAS protocol targeted the hand area of M1, and only 20 out of the 30 burn participants had a burn affecting their hand. It is possible that the effect of a burn injury on LTP-like plasticity is restricted to the representation of the burned body area in M1 (although Whife et al. [23] also targeted the hand area of M1 in a heterogenous sample in terms of burn location, demonstrating expected results). Future research should examine both LTP- and LTD-like plasticity in a homogeneous burn injury population across the acute, subacute, and chronic recovery phases to comprehensively characterise the effects of burn injury on neuroplasticity.

PAS induced an increase in LICI immediately and 15 minutes following PAS in both former burns patients and non-injured controls. PAS did not induce any change in SICI; therefore, the current findings suggest that PAS selectively modulates GABA_B_-mediated inhibition. This is consistent with previous research showing PAS-induced increase in LICI in younger adults [64]. Though it is important to note that one previous study found no change in LICI following PAS in healthy adults when using stimulation parameters comparable to the current study [65], suggesting changes in LICI following PAS might show high inter-individual response variability similar to the changes in MEP amplitude following PAS. Taken together, these findings suggest the PAS-induced increase in GABA_B_-mediated inhibition is typical in former burn patients 1–3 years following burn injuries. It is worth noting, however, that the functional significance of the PAS-induced increase in GABA_B_-mediated inhibition remains uncertain; while PAS-induced LICI modulation may reflect adaptive inhibitory capacity within M1, further research is needed to determine how such neurophysiological effects relate to recovery of motor function following burn injury.

### No Difference in SICI or LICI Between Former Burn Patients and Non-injured Control Participants

Although former burn patients (1–3 years post-minor injury) had smaller and more variable SICI ratios compared to controls (Figure 4), no significant difference in SICI was observed between the two groups. Pharmacological evidence suggests SICI is mediated by GABA_A_ receptor activity [66, 67]. Therefore, the study findings suggest that there was no change in GABA_A_ receptor activity that persists 1–3 years following a burn injury. While no previous research has examined GABAergic inhibition in the chronic phase following burn injury, one study found no difference in SICI between burn patients at 6 and 12 weeks post-injury compared to controls [34]. Taken together, these findings suggest that, at the group-level, there is no change in intracortical inhibition mediated by GABA_A_ receptor activity in the subacute or chronic recovery phases following burn injury.

Similar to the SICI results, although former burn patients (1–3 years post-minor injury) had smaller and more variable LICI ratios compared to controls, no significant group difference was observed (Figure 4). Given that LICI is mediated by GABA_B_ receptor activity [31], the current findings suggest no persistent change in GABA_B_ receptor mediated inhibition 1–3 years following burn injury. Although this is the first study to examine LICI in burn populations, previous research has provided preliminary evidence that the cSP—mediated by both GABA_A_ and GABA_B_ receptor activity—is reduced in individuals with upper limb burns sustained less than two years post-injury compared to controls [33]. It is important to note, however, that the subgroup analyses from previous research showing reduced cSP [33] was performed on a small sample (n=13) and must be interpreted cautiously. Notwithstanding this caution, it is possible that burn injury affects the active modulation of GABAergic inhibition, and SICI and LICI measured in the resting muscle (as was done in the current study) are not sufficiently sensitive to show burn-related changes in inhibition. Future research should measure SICI and LICI during voluntary muscle activation to determine whether burn injury affects the use-dependent modulation of SICI or LICI.

Furthermore, it should be noted that, to date, SICI and LICI have only been measured using a single conditioning stimulus intensity. Both SICI and LICI are sensitive to conditioning stimulus intensity, resulting in U-shaped curves when the amount of inhibition is observed as a function of increasing conditioning stimulus intensity [68–71]. The descending limbs of SICI and LICI recruitment curves, where inhibition increases with increasing stimulation intensity, represent the progressive recruitment of populations of inhibitory interneurons that mediate these processes [72]. It is important for future research to measure SICI and LICI at a range of conditioning stimulus intensities to determine whether the recruitment of populations of inhibitory interneurons is affected by burn injury.

### Greater Intracortical Inhibition is Associated with Better Bilateral Manual Dexterity in Former Burn Patients

Despite no group-level differences in SICI between former burn patients (1–3 years post-minor injury) and non-injured controls, a robust association was observed between SICI and bilateral manual dexterity in burn participants but not in controls (Figure 5). Specifically, in burn participants, greater SICI was linked to better performance on the simple bilateral subtest of the Purdue Pegboard. This suggests that, despite no significant group-level differences in SICI between burn participants and controls, the large variation in SICI magnitude among burn participants is likely functionally important, as SICI magnitude 1–3 years post-burn injury is associated with manual dexterity. Previous research supports the functional importance of SICI in the preparation and execution of dexterous movement: SICI is reduced before and during muscle contraction [73–76], and the reduction in SICI during muscle contraction is greater during precision grasps than isometric contractions [77]. Similarly, during movement, healthy participants showed reduced SICI responses in the synergistic muscles directly involved in the task (e.g., FDI during an index finger flexion task) and increased SICI in surrounding muscles (e.g., abductor pollicis brevis) that were not engaged in the task [78]. This pattern suggests that intracortical inhibition helps suppress the activation of muscles unrelated to the task—referred to as surround inhibition—potentially facilitating more efficient and controlled manual dexterity [79]. Additionally, in older adults with declining manual dexterity, a shift from inhibition to facilitation (or atypical facilitation) is associated with poorer manual dexterity [51]. In the current sample, six burn participants showed atypical facilitation (ratio exceeding 1.5), and these individuals generally performed worse on the manual dexterity tasks. The clinical characteristics of the atypical facilitation subgroup were similar to those of the entire sample, though they tended to be older; however, this study was not powered adequately to interpret subgroup results.

Similar to the associations between SICI and manual dexterity, despite no significant group-level differences in LICI between former burn patients (1–3 years post-minor injury) and non-injured controls, the wide variability in LICI magnitude among burn participants is likely functionally important. In this study, LICI magnitude was significantly associated with bilateral motor performance, with greater LICI linked to better performance on the simple bilateral and assembly pegboard subtests (Figure 6). Notably, this pattern contrasts with that observed in controls, where greater LICI was associated with poorer motor performance. Although the functional role of LICI is less well understood than that of SICI, there are notable similarities. Like SICI, LICI is reduced during voluntary contraction [80], and the reduction in LICI during muscle contraction is more pronounced during a precision grasp compared to an isometric contraction. [77]. Taken together, these findings suggest that LICI plays a functional role in upper limb control and that alterations in LICI may contribute to the reduction in motor capacity observed in former burn patients. Importantly however, the correlation analyses were exploratory, examining associations between intracortical inhibition in M1 and bilateral manual dexterity. Further research is needed to determine whether these neural mechanisms play a causal role in the lasting decline in motor function observed in individuals with a history of burn injury.

### No Difference in Self-reported Health and Well-being Between Former Burn Patients and Controls

Despite group-level differences in bilateral assembly subtest performance between former burn patients (1–3 years post-minor injury) and non-injured controls, overall self-reported health and well-being appear comparable between the two groups. These findings suggest that the reduced motor performance detected through the Purdue Pegboard task does not negatively impact the perceived well-being of individuals with a history of burn injury, particularly those with minor burns (<10% TBSA) who were discharged from the hospital years ago.

### Limitations and Future Directions

This study has several limitations and provides suggestions for future research that should be considered when interpreting the findings:

- While PAS was chosen to assess LTP-like plasticity due to its role in sensory-motor integration [48] and effectiveness in increasing MEP amplitude [45], literature on PAS-induced changes in MEP amplitude is inconsistent, with some studies showing increases [for example see: 48] and others showing no effect [for example see: 81]. Even when increases occur, response variability is high, with only ∼50% of participants responding as expected [82]. Two factors might have contributed to the PAS response variability in the current study: (1) standardised intervals may not account for individual differences in conduction time and result in suboptimal timing for plasticity induction [80]; (2) administering PAS at varied times of the day could lead to PAS response variability due to circadian fluctuations [84, 85].
- Significant associations between intracortical inhibition in the hand representation of M1 and manual dexterity suggest that, despite variability in burn location, changes in intracortical inhibitory excitability may reflect broader motor impairments. However, given the heterogeneity in burn injury location, it remains unclear whether burn injury specifically affects the cortical representation of the burn-affected body part or has more widespread effects on the motor cortex.
- The current study only examined excitability in the hemisphere contralateral to the burn injury; it remains unclear whether compensatory changes occur in the ipsilateral hemisphere. Future research should investigate corticospinal excitability and intracortical inhibition in cortical representations of burn-affected, adjacent, and distant body parts in both hemispheres, as well as their associations with functional outcomes.
- In the current study, unilateral subtests were categorised as dominant and non-dominant rather than most-affected and least-affected, as hand dominance influences manual dexterity [86]. Figure 2 (panels A and B) highlights the most-affected and least-affected sides relative to hand dominance, however, dexterity was not analysed based on these classifications due to insufficient statistical power for subgrouping. Future research should aim for a balanced sample of burn injury participants, ensuring equal representation of injured limb and hand dominance when assessing manual dexterity.
- Correlation analyses focused on bilateral pegboard subtests since TMS was conducted on one hemisphere, allowing inclusion of the full sample. However, bilateral pegboard performance depends on both sides of the body, limiting the explanatory power of these correlations. Future research should assess motor cortex and corticospinal function in both hemispheres to enable comprehensive correlation analyses across all pegboard subtests.
- Due to the cross-sectional nature of this study, it is not possible to determine whether former burn patients had poor manual dexterity prior to injury. The associations between baseline intracortical inhibition (SICI and LICI) and bilateral manual dexterity in burn participants indicate that greater inhibition is linked to better dexterity—a pattern not observed in controls. This suggests that pre-injury differences in manual dexterity are unlikely to fully account for these findings.
- While SF36 self-reported pain scores showed no significant differences between former burn patients (1–3 years post-minor injury) and non-injured controls, chronic pain can modulate cortical excitability through sustained activation of nociceptive pathways [87, 88]. Additionally, persistent pain has been shown to alter the neural representation of the injured body area in M1, potentially impacting motor function and neuroplasticity [89].
- Both pre-injury and post-injury medications, including pain relief and antidepressants [90], which act on the central nervous system, as well as surgical treatment [91] received by five burn participants in the current sample, may have influenced the observed findings. Future studies should account for these potential confounders by stratifying participants based on pain or medication use or incorporating these factors as covariates in analyses.

### Conclusions

Former burn patients (1–3 years post-minor injury) demonstrated poorer performance on the bilateral assembly subtest compared to non-injured controls. Exploratory analyses further revealed associations between bilateral pegboard performance and intracortical inhibition within the hand representation of M1, underscoring its functional relevance and suggesting that burn injury may influence inhibitory mechanisms. These findings suggest that measures of intracortical inhibition may offer valuable insights into motor function long after burn injury. However, further research is needed to determine whether these neural mechanisms play a causal role in motor rehabilitation and represent a therapeutic opportunity post-burn injury.

## Data Availability

The primary data are available via https://osf.io/3xfkp/?view_only=48fa8a1f1b1c473d9e7f4748f61ecaf0

## Disclosure statement

During the preparation of this work, the author(s) used ChatGPT (OpenAI) and Microsoft CoPilot to improve the readability of the manuscript. After using these tools, the author(s) reviewed and edited the content as needed and take(s) full responsibility for the content of the publication.

## Funding

This project was funded by a National Health and Medical Research Ideas Grant (GNT2004107) and the Fiona Wood Foundation. Ann-Maree Vallence was supported by an Australian Research Council Discovery Early Career Researcher Award (DE190100694).

## Conflict of Interest

The authors declare no conflicts of interest related to this manuscript.

## Notes

### Competing Interest Statement

The authors have declared no competing interest.

### Author Declarations

The experimental protocol was performed in accordance with the Declaration of Helsinki and was approved by the Murdoch University (2021/047) and South Metropolitan Health Service Human Research Ethics Committee (RGS0000004279).

