## Supplementary Materials for "Characterising Manual Dexterity, Motor Cortex Neuroplasticity and Intracortical Inhibition Long After Burn Injury"

**1.0	Full Sample Analysed**

1.1	Surveys (Former burn patients: n=30; Closely matched controls: n=29)

*1.1.1	SF36 domain scores for role limitations due to physical health in former burn patients (1–3 years post-minor injury) and controls*

*** Insert Figure S1 Here ***

Figure S1: SF36 domain scores for role limitations due to physical health in former burn patients (1-3 years post-minor injury) and controls. These results suggest that former burn patients experience role limitations due to physical health that are not significantly different from those of non-injured controls. However, a ceiling effect was present and should be considered when interpreting these results.

*1.1.2	Burn-specific Health Scale-Brief (BSHS-B)*

Figure S2 highlights BSHS-B domain scores for physical (left) and generic (right) scores in former burn patients 1–3 years post-minor injury. Control participants did not complete the BSHS-B; therefore, the burn injury sample was compared to normative mean scores from the general population [1]. Two domain scores were calculated from the BSHS-B: the generic score, reflecting psychological and social well-being, and the physical score, representing physical functional abilities. The normative mean scores were 71.3 for the generic domain and 34.8 for the physical domain [1]. The domain scores for former burn patients were 77.1 ± 6.6 for the generic score and 35.6 ± 1.1 for the physical score. Bootstrapped one-sample t-tests revealed that burn participants scored significantly higher than normative data for both the generic score (95% CI: 74.53 to 79.17, *p* < 0.001) and physical score (95% CI: 35.13 to 35.90, *p* = 0.001), suggesting better psychological, social, and physical outcomes compared to general population norms. One possible consideration when interpreting this finding is that those who choose to participate in research may be more proactive in their recovery. Although qualitative research suggests that individuals with burn injuries often struggle to adhere to exercise prescriptions after injury [2], participants in this study may have a more positive attitude toward recovery and be more inclined to adopt healthier lifestyles, including increased physical activity as part of their rehabilitation or ongoing health management [3]. It is again important to note the likely ceiling effects of the physical score, with 24 burn participants recording a maximum score of 36, meaning narrow confidence intervals could potentially underestimate uncertainty.

*** Insert Figure S2 Here ***

Figure S2: BSHS-B domain scores for physical (left) and generic (right) scores in former burn patients 1-3 years post-minor injury. The dashed line represents the normative mean score for the general population as reported in the literature (1). Burn participants scored significantly higher than the general population norms, indicating better psychological, social, and physical outcomes compared to the general population. However, a ceiling effect was present and should be considered when interpreting the physical score results.

*1.1.3	painDETECT*

Missing values in the painDETECT survey (n=1, control participant who did not submit the survey) were handled by excluding incomplete cases from the analysis. The total score for burn participants and controls were 3.3 ± 5.2 and 3.1 ± 5.6, respectively. Bootstrapped t-tests revealed no significant difference between groups for self-reported pain (95% CI: -2.95 to 3.07, *p* = 0.902). This suggests that former burn patients (1–3 years post-minor injury) and non-injured controls report similar levels of pain, as measured by the painDETECT survey. The absence of persistent pain experienced by the burn participants may be attributed to the relatively small size of their injuries. Importantly, 19 burn participants and 17 controls recorded the minimum score of zero, indicating a floor effect. This diminishes variability in the data, which may affect the reliability of bootstrap resampling and lead to an underestimation of uncertainty in the findings.

*1.1.4	Patient Observed Scar Assessment Scale (POSAS) – Version 2.0*

Control participants did not complete the POSAS; therefore, the burn injury sample was compared to the mean score reported in another burn cohort with non-severe injuries, collected 28 months post-injury [4]. This comparison aimed to determine if our sample exhibited similar scores to a comparable, homogenous cohort. The hypothesised total mean score was 2.2 [4]. The score for burn participants was 2.1 ± 1.4. Bootstrapped one-sample t-tests revealed no significant difference in perceived scar scores (95% CI: 1.56 to 2.64, *p* = 0.707), suggesting that the former burn patients (1–3 years post-minor injury) had similar scar perceptions to another non-severe burns cohort with injuries sustained around the same time. Importantly, 12 participants recorded the minimum possible score of 1, indicating a floor effect. This reduces variability in the data, which could impact the reliability of bootstrap resampling and potentially underestimate uncertainty in the results.

*1.1.5	QuickDASH*

Missing values in the QuickDASH survey (n=1, control participant who did not submit the survey) were managed by excluding incomplete cases from the analysis. The total score for burn participants and controls were 29.1 ± 6.8 and 30.2 ± 7.1, respectively. Bootstrapped t-tests showed no significant difference between groups for self-reported upper-body function (95% CI: -4.69 to 2.20, *p* = 0.587), indicating comparable upper-body function between former burn patients (1–3 years post-minor injury) and controls. Notably, 18 burn participants and 10 controls achieved the lowest possible score of 25, indicating the presence of a floor effect. This reduces data variability, which could impact the reliability of bootstrap resampling and result in an underestimation of uncertainty in the outcomes.

*1.1.6	Lower Limb Functional Index (LLFI)*

Missing data in the LLFI survey (n=1, a control participant who did not submit the survey) were handled by excluding incomplete cases from the analysis. The final sample included nine former burn patients with lower-body injuries and 29 non-injured controls, all of whom completed the LLFI survey. The results reflect the total score from the first part of the survey. The total score for burn participants and controls were 1.9 ± 2.5 and 0.6 ± 1.1, respectively. Bootstrapped t-tests revealed no significant difference in self-reported lower-body function between groups (95% CI: -0.20. to 3.00, *p* = 0.501), suggesting comparable lower-body function between former burn patients (1–3 years post-minor injury) and controls. It should be mentioned that non-parametric bootstrapping is a robust technique, particularly for handling unequal sample sizes, as it makes minimal assumptions about the data’s distribution. However, the imbalance in group sizes presents specific challenges. The smaller sample size may reduce variability, leading to biased resampling distributions and narrower confidence intervals. Additionally, the smaller group may not fully represent the population, which could further impact the robustness of the bootstrap estimates. While the method remains appropriate, these limitations should be acknowledged when interpreting the findings, particularly for the smaller group.

1.2	Sensory Function

For former burn patients, sensory testing sites were marked on the skin using a surgical marker, approximately 3 cm from the scar. Testing was conducted at the following locations: above the scar on the most-affected side, the corresponding area on the least-affected side, below the scar on the most-affected side, and the corresponding area on the least-affected side. Sensory testing was conducted in a randomised order at each testing site.

*1.2.1	Pressure Pain Threshold*

A pressure algometer (FDX, Wagner Instruments, Greenwich, CT) with an 8-mm hemispheric rubber tip was used to measure pressure pain thresholds (Kg) at designated sensory testing sites. In addition, for both groups, measurements were taken at three standardised locations: (1) the forehead, where a familiarisation trial was conducted at the start; (2) the back of the hand on the most-affected side for burn participants and the dominant hand for controls; and (3) the back of the hand on the least-affected side for burn participants and the non-dominant hand for controls.

*1.2.1.1	Comparing Above and Below Scar Locations Within Burn Injury Participants*

Figure S3 shows the difference in pressure pain threshold scores between the most affected-above site and the least affected-above site in former burn patients (1–3 years post-minor injury). Missing values in the pressure pain threshold test were managed by excluding incomplete cases: 24 participants were included in the comparison between the most affected-above and least affected-above sites, while 22 participants were included in the comparison between the most affected-below and least affected-below sites. Above scar tests were not undertaken if they were too close to sensitive areas such as the genital region or face, and below scar tests were also not undertaken if the injury was located at the end of a limb. The scores for burn participants were as follows: 4.1 ± 2.1 for the most affected-above, 4.9 ± 2.2 for the least affected-above, 4.4 ± 2.5 for the most affected-below, and 4.5 ± 2.3 for the least affected-below. Bootstrapped paired t-tests revealed no significant between-limb differences in pressure pain threshold at above (95% CI: -1.57 to -0.12, *p* = 1.000) and below scar locations (95% CI: -1.26 to 0.62, *p* = 1.000). These findings suggest no differences in pain sensitivity around the injury site compared to the homologous sites on the least-affected side.

*** Insert Figure S3 Here ***

Figure S3: Difference in pressure pain threshold scores (Kg) between the most affected-above site and the least affected-above site in former burn patients with a minor injury. These findings suggest that there are no pain sensitivity differences above the scar when compared to the homologous site on the least-affected side.

*1.2.1.2	Comparing Standardised Testing Locations Between Burn and Control Groups, and Self-reported Pain Attributed to the Burn Injury*

Missing values in the pressure pain threshold test for the dominant hand, non-dominant hand, and forehead were addressed by excluding incomplete cases. Testing was omitted at certain sites in burn participants due to overlap with above-below scar locations or at the participant's request. This resulted in sample sizes of 21 burn participants and 30 controls for the dominant hand, 20 burn participants and 30 controls for the non-dominant hand, and 24 burn participants and 30 controls for the forehead. The scores for former burn patients were as follows: 3.4 ± 1.1 for the dominant hand, 3.6 ± 1.2 for the non-dominant hand, and 2.1 ± 0.7 for the forehead. For the controls the scores were as follows: 3.1 ± 1.6 for the dominant hand, 3.3 ± 1.3 for the non-dominant hand, and 2.1 ± 0.7 for the forehead. Bootstrapped t-tests revealed no significant differences in pressure pain threshold between groups for the dominant hand (95% CI: -0.50 to 1.01, *p* = 1.000), non-dominant hand (95% CI: -0.39 to 1.06, *p* = 1.000), and forehead (95% CI: -0.37 to 0.41, *p* = 1.000). These results suggest that pressure pain threshold measurements at non-injured control sites are similar between former burn patients (1–3 years post-minor injury) and controls.

*1.2.2	Visual Analogue Pain Scale*

Self-reported pain, assessed using a visual analogue scale (0-100) specifically attributed to the burn injury, was 2.9 ± 6.4 (95% CI: 0.80 to 5.14, *p* < 0.001). This indicates that former burn patients experienced a low level of pain, significantly different from zero, suggesting mild persistent pain associated with the injury. However, it is worth noting that 23 burn participants recorded the minimum possible score of zero, highlighting a floor effect. This limits variability in the data, potentially affecting the reliability of bootstrap resampling and leading to an underestimation of uncertainty in the results.

*1.2.3	Semmes-Weinstein Monofilament Test (SWMT) Within Former Burn Patients*

A set of six sensory monofilaments (Touch-Test Sensory Monofilaments, North Coast Medical, USA) was utilised to assess touch sensitivity. Missing values in the Semmes-Weinstein Monofilament Test were addressed by excluding incomplete cases. Above scar tests were omitted if they were too close to sensitive areas, such as the genital region or face, while below scar tests were also excluded if the injury was located at the end of a limb. As a result, the sample sizes were 28 for comparisons between the most affected-above and least affected-above sites, and 26 for comparisons between the most affected-below and least affected-below sites. The scores for former burn patients were as follows: 3.2 ± 0.5 for the most affected-above, 3.2 ± 0.5 for the least affected-above, 3.2 ± 0.8 for the most affected-below, and 3.2 ± 0.5 for the least affected-below. Bootstrapped paired t-tests revealed no significant differences in Semmes-Weinstein Monofilament Test between the most affected and least affected sides at both above (95% CI: -0.16 to 0.27, *p* = 1.000) and below scar locations (95% CI: -0.23 to 0.11, *p* = 1.000). These findings suggest no significant, measurable differences in touch sensitivity between the most and least affected sides at either location. Future research should consider testing at standard locations, such as the fingertips, to compare touch sensitivity between burn participants and non-injured controls.

*1.2.4	Neuropen Within Former Burn Patients*

Table S1 presents the Neuropen mean and standard deviation scores for former burn patients (1–3 years post-minor injury), along with statistical significance results. Figure S4 highlights the difference in Neuropen Dull scores, following both 1 trial (left) and 5 trials (right), between the most affected-above site and the least affected-above site in burn participants with a minor injury. Participants assessed the sharpness of both a filament and Neurotip (Neuropen, Owen Mumford, UK) under two conditions: (1) a single application of the stimulus, and (2) five repeated applications of the stimulus. Sharpness was rated using a verbal rating scale ranging from 0 (not sharp) to 10 (extremely sharp). Missing values in the Neuropen tests were handled by excluding incomplete cases, resulting in a sample size of 28 for above scar site comparisons. For below scar sites, the sample sizes were 26 for the comparison of most and least affected dull (1-trial), and 27 for the comparisons of dull (5-trial) and sharp (1-trial and 5-trial) between most and least affected sites. Bootstrapped paired t-tests revealed no significant differences in Neuropen results for the comparisons between the most affected-above and least affected-above sites under dull and sharp conditions, nor between the most affected-below and least affected-below sites (Table S1). These findings suggest that touch sensitivity does not vary significantly around the injury site between the most and least affected sides. Notably, 19 data points for the most affected-above dull (1-trial), 16 for the most affected-above dull (5-trial), 21 for the least affected-above dull (1-trial), and 17 for the least affected-above dull (5-trial) had the minimum possible score of zero, indicating a floor effect. This limited variability in the data could affect the reliability of bootstrap resampling and potentially lead to an underestimation of uncertainty in the results.

*** Insert Figure S4 Here ***

Figure S4: Difference in Neuropen Dull scores, following both 1 trial (left) and 5 trials (right), between the most affected-above site and the least affected-above site in former burn patients with a minor injury. These findings suggest that touch sensitivity does not vary significantly around the injury site between the most and least affected sides. However, a floor effect was present and should be considered when interpreting the results.

*** Insert Table S1 Here ***

*1.2.5	Brush Perception Test Within Former Burn Patients*

Touch perception was evaluated by gently sweeping a lightweight paintbrush back and forth over the designated sensory testing sites. Missing values in the brush perception tests were handled by excluding incomplete cases. Certain sites were excluded from testing due to the injury's location or at the participant's request. The brush perception test was used to assess sensory perception at above and below scar sites, with responses categorised as “Normal,” “Painful,” “Weaker than expected,” “Stronger than expected,” or “Abnormal.” The distribution of responses across these categories is as follows: at most affected-above, 100% of responses were categorised as "Normal" (29/29). At most affected-below, 92.9% of responses were "Normal" (26/28), and 7.1% were "Weaker" (2/28). For least affected-above, 89.7% were "Normal" (26/29), 6.9% were "Weaker" (2/29), and 3.5% were "Stronger" (1/29). Finally, for least affected-below, 96.4% of responses were categorised as "Normal" (27/28), and 3.6% as "Abnormal" (1/28). These results suggest that light touch perception in former burn patients was predominantly normal, with only a small proportion of responses categorised as weaker or abnormal. This suggests that light touch sensitivity is generally intact when comparing the most-affected side to the least-affected side at both above and below scar sites around the injury.

1.3	Grip Strength

*1.3.1	Comparing Dominant Hand and Non-dominant Hand Grip Strength Between Burn and Controls Groups*

Figure S5 shows the comparison of grip strength in the dominant and non-dominant hands between former burn patients (1–3 years post-minor injury) and non-injured controls. Grip strength was assessed in both hands using a handgrip force dynamometer (TTM Advanced Hand Dynamometer, Japan). For the burn participants the scores were 43.4 ± 13.0 for the dominant hand and 41.1 ± 12.6 for the non-dominant hand. For the controls the scores were 39.4 ± 11.1 for the dominant hand and 37.6 ± 11.0 for the non-dominant hand. Bootstrapped t-tests revealed no significant differences in grip strength between groups for the dominant hand (95% CI: -2.09 to 10.63, *p* = 1.000) and non-dominant hand (95% CI: -2.25 to 8.91, *p* = 1.000). These findings suggest that grip strength was comparable between former burn patients and controls for both hands.

*** Insert Figure S5 ***

Figure S5: Comparison of grip strength in the dominant and non-dominant hands between former burn patients (1-3 years post-minor injury) and non-injured controls. The results indicate no significant differences in grip strength between the two groups for either hand.

1.4	Neurophysiological Outcomes

*1.4.1	SICI and LICI Following PAS in Former Burn Patients and Controls – Uncropped Figure*

Short-interval intracortical inhibition (SICI) and long-interval intracortical inhibition (LICI) ratios following PAS, comparing former burn patients (1–3 years after minor injuries) with non-injured controls.

*** Insert Figure S6 Here ***

Figure S6 (uncropped figure). Short-interval intracortical inhibition (SICI) and long-interval intracortical inhibition (LICI) ratios following PAS for former burn patients (1-3 years post-minor injury) (panels A and C, respectively) and non-injured controls (panels B and D, respectively). Points that lie beyond the whiskers are shown as filled circles, indicating aggregated-level outliers.

*1.4.2	Associations Between MEP Amplitude, SICI, and LICI Changes Following PAS and Bilateral Pegboard Performance*

Figure S7 shows associations between changes in MEP amplitude following PAS (PRE vs. 0-min POST-PAS) and bilateral pegboard performance. Missing values for the MEP, SICI, and LICI changes and pegboard correlation analyses (n=3, burn injury participants who did not complete post-PAS testing due to discomfort from peripheral stimulation) were managed by excluding incomplete cases from the analysis. Bootstrapped correlation analyses were conducted to evaluate these relationships in both the burn and control groups. For the burn group, no significant correlations were found between PAS-induced neuroplasticity (MEP change) and the bilateral pegboard subtests. Specifically, the correlations were *r* = -0.09 (95% CI: -0.44 to 0.32) for the simple bilateral subtest (Figure S7A) and *r* = -0.12 (95% CI: -0.49 to 0.27) for the assembly subtest (Figure S7B). Similarly, no significant correlations were observed in the control group. The correlations were *r* = -0.31 (95% CI: -0.63 to 0.10) for the simple bilateral subtest (Figure S7A) and *r* = -0.10 (95% CI: -0.43 to 0.25) for the assembly subtest (Figure S7B). Additionally, bootstrapped analyses comparing the relationships between the burn and control groups showed no significant differences. These results indicate that changes in MEP amplitude following PAS do not significantly correlate with bilateral manual dexterity performance on the Purdue Pegboard in either the former burn patient or control groups.

*** Insert Figure S7 Here ***

Figure S7. The associations between changes in MEP amplitude following PAS (PRE vs. 0-min POST-PAS) and bilateral pegboard performance (panel A: simple bilateral, panel B: assembly) in former burn patients (1–3 years post-minor injury) (blue circle symbols) and controls (grey triangle symbols).

Figure S8 shows associations between changes in SICI following PAS (PRE vs. 0-min POST-PAS) and bilateral pegboard performance. No significant correlations were found between PAS-induced neuroplasticity (SICI change) and the bilateral pegboard subtests in either group. In the burns group, the correlations were *r* = -0.18 (95% CI: -0.48 to 0.19) for the simple bilateral subtest (Figure S8A) and *r* = -0.16 (95% CI: -0.40 to 0.05) for the assembly subtest (Figure S8B). For the control group, the correlations were *r* = -0.37 (95% CI: -0.63 to 0.01) for the simple bilateral subtest (Figure S8A) and *r* = -0.04 (95% CI: -0.37 to 0.27) for the assembly subtest (Figure S8B). This suggests that changes in SICI following PAS do not have a strong impact on bilateral pegboard performance in either group.

*** Insert Figure S8 ***

Figure S8. The associations between changes in SICI following PAS (PRE vs. 0-min POST-PAS) and bilateral pegboard performance (panel A: simple bilateral, panel B: assembly) in former burn patients (1–3 years post-minor injury) (blue circle symbols) and controls (grey triangle symbols).

Figure S9 shows associations between changes in LICI following PAS (PRE vs. 0-min POST-PAS) and bilateral pegboard performance. No significant correlations were found between PAS-induced neuroplasticity (LICI change) and the bilateral pegboard subtests in either group. In the burns group, the correlations were *r* = 0.02 (95% CI: -0.32 to 0.34) for the simple bilateral subtest (Figure S9A) and *r* = 0.03 (95% CI: -0.31 to 0.31) for the assembly subtest (Figure S9B). For the control group, the correlations were *r* = -0.21 (95% CI: -0.52 to 0.14) for the simple bilateral subtest (Figure S9A) and *r* = -0.38 (95% CI: -0.72 to 0.06) for the assembly subtest (Figure S9B). Additionally, bootstrapped analyses comparing the relationships between the burn and control groups showed no significant differences. These findings indicate that changes in LICI following PAS are not strongly associated with bilateral pegboard performance in either the burn or control group, and no significant differences were observed between the groups.

*** Insert Figure S9 ***

Figure S9. The associations between changes in LICI following PAS (PRE vs. 0-min POST-PAS) and bilateral pegboard performance (panel A: simple bilateral, panel B: assembly) in former burn patients (1–3 years post-minor injury) (blue circle symbols) and controls (grey triangle symbols).

***2.0	Former Burn Patients with Upper Limb Injuries (n=21)***

2.1	Summary of Clinical Characteristics for Former Burn Patients with Upper Limb Injuries (n=21) and Descriptives of Non-injured Controls (n=21)

A subgroup analysis of former burn patients with upper limb injuries (n=21) was performed to evaluate its impact on TMS outcomes and pegboard performance. Upper limb injury was defined as any reported trauma involving the upper arm, forearm, or hand. The clinical profile of the burn participants (1–3 years post-injury) with upper limb injuries included an average time since injury of 85 ± 18 weeks, a mean TBSA affected of 0.83 ± 1.13%, and the following distribution of burn types: contact (n=7), scald (n=7), flame (n=5), and electrical (n=2). Burn depth was categorised as superficial (n=6), partial-thickness (n=10), deep (n=4), and full-thickness (n=1). A weighted matching algorithm was employed to create a control subgroup closely aligned with burn participants who had upper limb injuries. Matching criteria included sex (burn group: females = 8; control group: females = 8), age (burn group: 40.8 ± 13.3 years; control group: 41.1 ± 13.6 years) and the dominant hemisphere tested with TMS (burn group: n=17; control group: n=17).

2.2	Pegboard Performance Differences Between Former Burn Patients with Upper Limb Injury and Non-injured Controls

Figure S10 displays scatterplots of pegboard performance across the four subtests comparing former burn patients (1–3 years post-minor injury) with upper-limb injuries and non-injured controls. The pegboard scores for burn participants were 14.0 ± 1.8 for the dominant hand, 13.5 ± 1.8 for the non-dominant hand, 10.2 ± 1.7 for the simple bilateral task, and 29.2 ± 4.5 for the assembly task. For non-injured controls, the scores were 15.1 ± 1.8 for the dominant hand, 13.8 ± 1.5 for the non-dominant hand, 11.3 ± 1.3 for the simple bilateral task, and 34.3 ± 7.2 for the assembly task. Bootstrapped t-tests showed that former burn patients demonstrated poorer performance compared to controls for the assembly subtest (95% CI: -8.60 to -1.61, *p* = 0.038, *d* = -0.85). However, no significant differences were observed between groups for the dominant hand subtest (95% CI: -2.21 to -0.03, *p* = 0.115, *d* = -0.60), the non-dominant hand subtest (95% CI: -1.25 to 0.75, *p* = 0.574, *d* = -0.18), or the simple bilateral subtest (95% CI: -2.06 to -0.34, *p* = 0.058, *d* = -0.75). Effect size differences across pegboard subtests were non-significant, indicating comparable group differences across all measures. The largest contrast—between assembly (*d* = -0.85) and non-dominant hand (*d* = -0.18)—did not reach statistical significance (95% CI: -1.47 to 0.10), suggesting that while the assembly task showed a trend toward a larger effect size (effect size difference = -0.68), the difference between tasks remains uncertain. These findings align with those reported in the full sample analysis. Notably, the smaller sample size in this subgroup may have contributed to increased variability, reducing statistical power and making it more challenging to detect a significant effect size difference between pegboard subtests.

*** Insert Figure S10 Here ***

Figure S10. Dominant hand, non-dominant hand, simple bilateral, and assembly pegboard performance for former burn patients (1-3 years post-minor injury) with upper limb injuries and non-injured controls. An asterisk indicates a statistically significant difference between groups.

2.3	Neurophysiological Outcomes

*2.3.1	Baseline TMS Intensities (RMT; SI_1mV_)*

For burn participants, the RMT and SI_1mV_ were 51.2 ± 8.0% (MSO) and 60.7 ± 10.1% (MSO), respectively. For the non-injured controls, the RMT was 49.0 ± 6.7% (MSO), and the SI_1mV_ was 57.4 ± 7.4% (MSO). Bootstrapped t-tests were conducted to compare the mean RMT and SI_1mV_ scores between former burn patients with upper limb injuries and their matched controls. There were no differences between groups for RMT (95% CI: -1.83 to 6.65, *p* = 0.484) or SI_1mV_ (95% CI: -1.58 to 8.40, *p* = 0.484). These findings align with those reported in the full sample analysis.

*2.3.2	MEP Amplitude, SICI, and LICI Following PAS*

Figure S11 shows MEP changes following PAS in former burn patients (1–3 years post-minor injury) with upper limb injuries compared to non-injured controls. Figure S12 shows SICI and LICI changes after PAS in former burn patients with upper limb injuries compared to non-injured controls. A generalised linear mixed model (GLMM) was used to evaluate changes in MEP amplitude, SICI, and LICI between burn participants who had an upper limb injury and matched controls. No significant effects were found for GROUP, χ2 (N = 42) = 0.62, *p* = 0.431, or TIME, χ2 (N = 42) = 0.28, *p* = 0.964. However, there were significant main effects for TMS TYPE, χ2 (N = 42) = 77.90, *p* < 0.001, and a TIME x TMS TYPE interaction, χ2 (N = 42) = 30.86, *p* < 0.001. No other significant interaction effects were detected, including those related to GROUP. Post hoc analyses revealed no significant changes in single-pulse MEP amplitude (*z* < 0.90, *p* = 1.000) or SICI (*z* < 1.80, *p* > 0.254) across TIME, but LICI significantly increased from PRE at 0-min POST-PAS (*z* = -4.02, *p* < 0.001) and 15-min POST-PAS (*z* = -2.58, *p* = 0.049). These findings are consistent with those observed in the full sample.

*** Insert Figure S11 Here ***

Figure S11. Single-pulse MEP amplitude following PAS in former burn patients (1-3 years post-minor injury) with upper limb injuries and their matched controls. The lower and upper hinges correspond to the 25th and 75th percentiles, respectively. Whiskers extend to values within 1.5 times the interquartile range (the difference between these percentiles) below the lower hinge and above the upper hinge. Data points beyond the whiskers are displayed as filled circles, representing trial-level outliers.

*** Insert Figure S12 Here ***

Figure S12. Short-interval intracortical inhibition (SICI) and long-interval intracortical inhibition (LICI) ratios following PAS in former burn patients (1-3 years post-minor injury) with upper limb injuries (panels A and C, respectively) and their controls (panels B and D). The y-axis of the LICI ratio plots has been cropped at 1.6 to highlight the boxplots.

*2.3.3	Baseline SICI and LICI Comparison Between Former Burn Patients with Upper Limb Injuries and Matched Controls*

A generalised linear mixed model (GLMM) was used to evaluate baseline differences in SICI and LICI between former burn patients (1-3 years post-minor injury) with upper limb injuries and their matched controls. There were no significant effects for GROUP, χ2 (N = 42) = 1.23, *p* = 0.266, and no significant GROUP x TMS TYPE interaction, χ2 (N = 42) = 2.52, *p* = 0.283. However, there was a significant main effect of TMS TYPE, χ2 (N = 42) = 88.50, *p* < 0.001: LICI ratios were numerically smaller than SICI ratios. These findings align with those observed in the full sample.

*2.3.4	Relationships Between Baseline SICI and LICI and Bilateral Pegboard Performance in Former Burn Patients with Upper Limb Injuries*

Figures S13 and S14 display scatterplots showing the association between the magnitude of baseline (PRE) SICI and LICI and bilateral pegboard performance in former burn patients (1–3 years post-minor injury) with upper limb injuries and controls. In the subgroup of burn participants with upper limb injuries, bootstrapped correlation analyses revealed negative relationships between baseline SICI and simple bilateral (*r* = -0.40, 95% CI: -0.72 to -0.06) and assembly performance (*r* = -0.35, 95% CI: -0.67 to -0.01), suggesting that greater baseline SICI is associated with better motor performance on these tests. The significant association with the assembly test is a novel finding that was not observed in the full sample. No significant associations were observed for the matched control group.

*** Insert Figure S13 Here ***

Figure S13. The relationships between the magnitude of SICI and pegboard performance (panel A: simple bilateral, panel B: assembly) in former burn patients (1-3 years post-minor injury) with upper limb injuries (blue circle symbols) and their matched controls (grey triangle symbols).

A significant negative correlations were observed between baseline LICI and simple bilateral pegboard performance (*r* = -0.51, 95% CI: -0.76 to -0.22) in former burn patients (1–3 years post-minor injury), indicating that greater baseline LICI is associated with better manual dexterity. This finding aligns with the results from the full sample, except that the full sample also showed a significant correlation with assembly performance. In the control group, the only significant correlation observed was with simple bilateral performance (*r* = 0.32, 95% CI: 0.03 to 0.62), consistent with the findings from the full sample analysis. Significant correlation differences were identified between burn participants with upper limb injuries and their matched controls for baseline LICI and simple bilateral performance (95% CI: -1.24 to -0.38) and assembly performance (95% CI: -1.19 to -0.20). These findings are consistent with the results from the full sample.

*** Insert Figure S14 Here ***

Figure S14. The relationships between the magnitude of LICI and pegboard performance (panel A: simple bilateral, panel B: assembly) in former burn patients (1-3 years post-minor injury) with upper limb injuries (blue circle symbols) and their matched controls (grey triangle symbols).

2.4	Self-reported Health and Well-being - Short Form-36 (SF36)

Table 2 in the primary manuscript presents the domain scores for the full sample. Bootstrapped t-tests revealed no significant differences between former burn patients (1–3 years post-minor injury) with upper limb injuries and non-injured controls across all SF36 domain scores. These results suggest that the overall health and well-being of burn participants with upper limb injuries are not significantly different from those of non-injured controls. These findings align with the results from the full sample.
