## Supplementary material for "Characterising Manual Dexterity, Motor Cortex Neuroplasticity and Intracortical Inhibition Long After Burn Injury": Table S1

Table S1: Neuropen mean and standard deviation scores in former burn patients (1–3 years post-minor injury), along with statistical significance results.

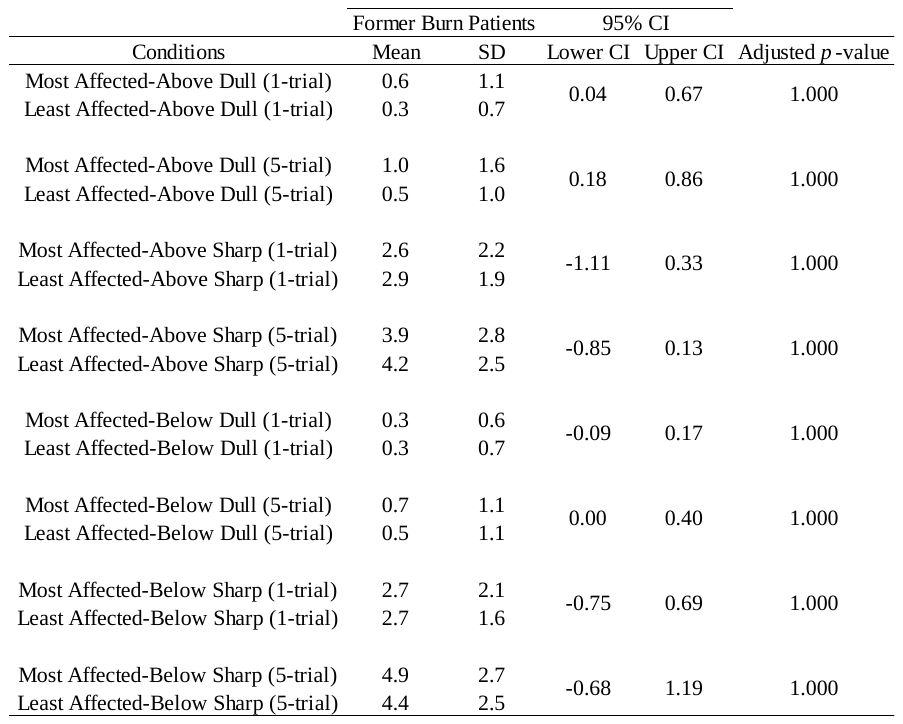

SD = standard deviation; CI = confidence interval
